# Explainable AI-based analysis of human pancreas sections identifies traits of type 2 diabetes

**DOI:** 10.1101/2024.10.23.24315937

**Authors:** L Klein, S Ziegler, F Gerst, Y Morgenroth, K Gotkowski, E Schöniger, M Heni, N Kipke, D Friedland, A Seiler, E Geibelt, H Yamazaki, HU Häring, S Wagner, S Nadalin, A Königsrainer, A L Mihaljević, D Hartmann, F Fend, D Aust, J Weitz, Jumpertz-von Schwartzenberg R, M Distler, K Maier-Hein, A L Birkenfeld, S Ullrich, P Jäger, F Isensee, M Solimena, R Wagner

**Affiliations:** IML Group, DKFZ, Heidelberg, Germany; Institute for Machine Learning, ETH Zürich, Zürich, Switzerland; Helmholtz Imaging, DKFZ, Heidelberg, Germany; Division of Medical Image Computing, DKFZ, Heidelberg, Germany; Institute for Diabetes Research and Metabolic Diseases of the Helmholtz Center Munich at the Eberhard Karls University of Tuebingen (IDM), Tuebingen, Germany; Internal Medicine IV, Endocrinology, Diabetology and Nephrology, University Hospital Tuebingen, Tuebingen, Germany; German Center for Diabetes Research (DZD e.V.) Neuherberg, Germany; Department of Molecular Diabetology, University Hospital Carl Gustav Carus, Medical Faculty, Technische Universität Dresden, Dresden, Germany; Paul Langerhans Institute Dresden (PLID) of the Helmholtz Center Munich at the University Hospital Carl Gustav Carus and Faculty of Medicine of the TU Dresden, Dresden, Germany; Internal Medicine I, Endocrinology and Diabetology, University Hospital Ulm, Ulm, Germany; Institute for Clinical Chemistry and Pathobiochemistry, Department for Diagnostic Laboratory Medicine, University Hospital Tübingen, Tübingen, Germany; Center for Molecular and Cellular Bioengineering at the Technische Universität Dresden Light Microscopy Facility (TUD CMCB LMF); Section of Clinical Epidemiology, Department of Community Medicine, Graduate School of Medicine, Kyoto University, Kyoto, Japan; Center for Innovative Research for Communities and Clinical Excellence (CiRC2LE), Fukushima Medical University, Fukushima, Japan; Department of General, Visceral and Transplant Surgery, University Hospital Tuebingen, Tuebingen, Germany; Institute of Pathology and Neuropathology and Comprehensive Cancer Center, University Hospital Tübingen, Tübingen, Germany; Department of Pathology, University Hospital Carl Gustav Carus, Medical Faculty, Technische Universität Dresden, Dresden, Germany; Department of Diabetes, Life Sciences & Medicine Cardiovascular Medicine & Sciences, Kings College London, London, United Kingdom; Department of Endocrinology and Diabetology, Medical Faculty and University Hospital Düsseldorf, Heinrich Heine University Düsseldorf, Düsseldorf, Germany; Institute for Clinical Diabetology, German Diabetes Center, Leibniz Center for Diabetes Research at Heinrich Heine University Düsseldorf, Düsseldorf, Germany; Pattern Analysis and Learning Group, Department of Radiation Oncology, Heidelberg University Hospital, Heidelberg, Germany

## Abstract

Type 2 diabetes (T2D) is a chronic disease currently affecting around 500 million people worldwide with often severe health consequences. Yet, histopathological analyses are still inadequate to infer the glycaemic state of a person based on morphological alterations linked to impaired insulin secretion and β-cell failure in T2D. Giga-pixel microscopy can capture subtle morphological changes, but data complexity exceeds human analysis capabilities. In response, we generated a dataset of pancreas whole-slide images with multiple chromogenic and multiplex fluorescent stainings and trained deep learning models to predict the T2D status. Using explainable AI, we made the learned relationships interpretable, quantified them as biomarkers, and assessed their association with T2D. Remarkably, the highest prediction performance was achieved by simultaneously focusing on islet α-and δ-cells and neuronal axons. Subtle alterations in the pancreatic tissue of T2D donors such as smaller islets, larger adipocyte clusters, altered islet-adipocyte proximity, and fibrotic patterns were also observed. Our innovative data-driven approach underpins key findings about pancreatic tissue alterations in T2D and provides novel targets for research.

## Introduction

Based on the current WHO classification of diabetes, over 90% of all persons with the condition fall into the category defined as type 2 diabetes (T2D). T2D is a major global health issue, affecting millions and placing a significant burden on healthcare systems (IDF Diabetes Atlas, 2021). Pathophysiologic drivers of T2D are insulin resistance and impaired insulin secretion, with dysfunction of pancreatic islet β-cells being the key feature. Despite extensive research, the exact etiology and nature of β-cell failure in the pathophysiology of T2D remain unclear. β-cells reside in specialized micro-organs, the islets of Langerhans, within the pancreas, but accessing human pancreatic tissue in living subjects is challenging due to anatomical constraints and the risks associated with pancreatic biopsy or tissue resection (Krogvold et al., 2014). Consequently, the identification of morphological islet changes associated with T2D has relied mainly on the histopathological analysis of specimens from deceased donors. Such studies pointed to several structural alterations such as amyloid deposition (Kahn et al., 1999; Westermark et al., 2011), reduced β-cell mass with decreased β-cell/α-cell ratio (Haataja et al., 2008; Weir et al., 2020), increased number of islet resident macrophages (Ehses et al., 2007; Richardson et al., 2009) and fibrosis (Deng et al., 2004; Hayden, 2007), but several of these findings remain controversial (Sempoux et al., 2001; Cohrs et al., 2020; Holman et al., 2020). Given the high degree of inter-individual variability, none of these traits is sufficient for pathologists to discriminate with a high degree of certainty whether a pancreatic specimen belongs to a subject with or without T2D. While such discriminative power would not have diagnostic relevance, it could pave the way to uncover yet unknown structural hallmarks of the disease and thereby provide further insight into its pathogenesis. This knowledge could in turn be leveraged to improve the prevention and treatment of diabetes.

Recent advances in Machine Learning (ML) have garnered significant attention in the field of computational pathology due to their ability to analyze large volumes of high-dimensional whole-slide images (WSIs) with efficacy comparable to expert pathologists (Bulten et al., 2022). For example, deep learning (DL) technologies are employed for prostate cancer grading (Bulten et al., 2022), cell segmentation (Shaban et al., 2024), gene expression prediction (Rahaman et al., 2023), and the discovery of new morphological biomarkers in breast cancer histopathology (Mandair et al., 2023). However, most DL applications in histopathology focus on oncological diseases, where distinct cancer cells are identifiable in tissue samples. In contrast, pancreatic tissue lacks clear quantifiable features that unequivocally differ between individuals with and without T2D.

Hence, our goal was the identification of histologic biomarkers allowing us to distinguish a pancreatic specimen from a patient with or without T2D and thus gain a better understanding of the disease. For that purpose, we exploited an imaging data-driven approach combining state-of-the-art DL with explainable artificial intelligence (XAI). While DL models could find patterns in data that are infeasible to detect by traditional approaches, XAI methods subsequently rendered the learned relationships human-understandable, revealing regions of interest (ROIs) associated with the presence of T2D. Given the limited feasibility of qualitative analyses of large amounts of XAI results, we quantified the attention to the ROIs and subsequently computed specific histologic biomarkers for the most important ones. At last, we analyzed these biomarkers in combination with clinical patient data using statistical models. Besides the integration of vast amounts of diverse WSIs, our approach offered the advantage that we did not bias ourselves to prior assumptions regarding T2D as the XAI application could also uncover unanticipated findings, leading to the formulation of new hypotheses about the condition.

## Results

### A unique dataset of chromogenic and fluorescence stained pancreas sections

DL models that can reliably distinguish patients with T2D from other patients are a prerequisite for gaining new insights into T2D through XAI methods. By uncovering the patterns used by the DL models for prediction, we aim for a better understanding of the disease.

To this end, we first collected a dataset by applying both single chromogenic and multiplex fluorescence techniques to immunostain pancreas sections from 100 metabolically phenotyped living donors with (n=35) or without (n=65) T2D who underwent pancreatectomy for various pancreatic disorders (Solimena et al., 2018; Gerst et al., 2017, 2018; Barovic et al., 2019; Gloyn et al., 2022) at two academic centers. Sections were immunostained for glucagon, insulin, and somatostatin as markers of islet α-, β- and δ-cells, respectively, as well as PECAM1 for endothelial cells to visualize islet vascularization, perilipin 1 for adipocytes to assess intra-pancreatic steatosis and tubulin beta 3 for neuronal axons. In the case of chromogenic stainings, each marker was detected individually in serial sections counterstained with hematoxylin. In the case of multiplex fluorescence stainings, serial sections were incubated with whole stainingsets, including DAPI and primary antibodies either against glucagon, somatostatin, and tubulin beta 3 (Stainingset 1), or against insulin, PECAM1, and perilipin 1 (Stainingset 2) (Fig. 1A and Method Section). Overall, eight images per patient were acquired. The dataset will be made publicly available alongside code for all following analyses.

**Figure 1.**
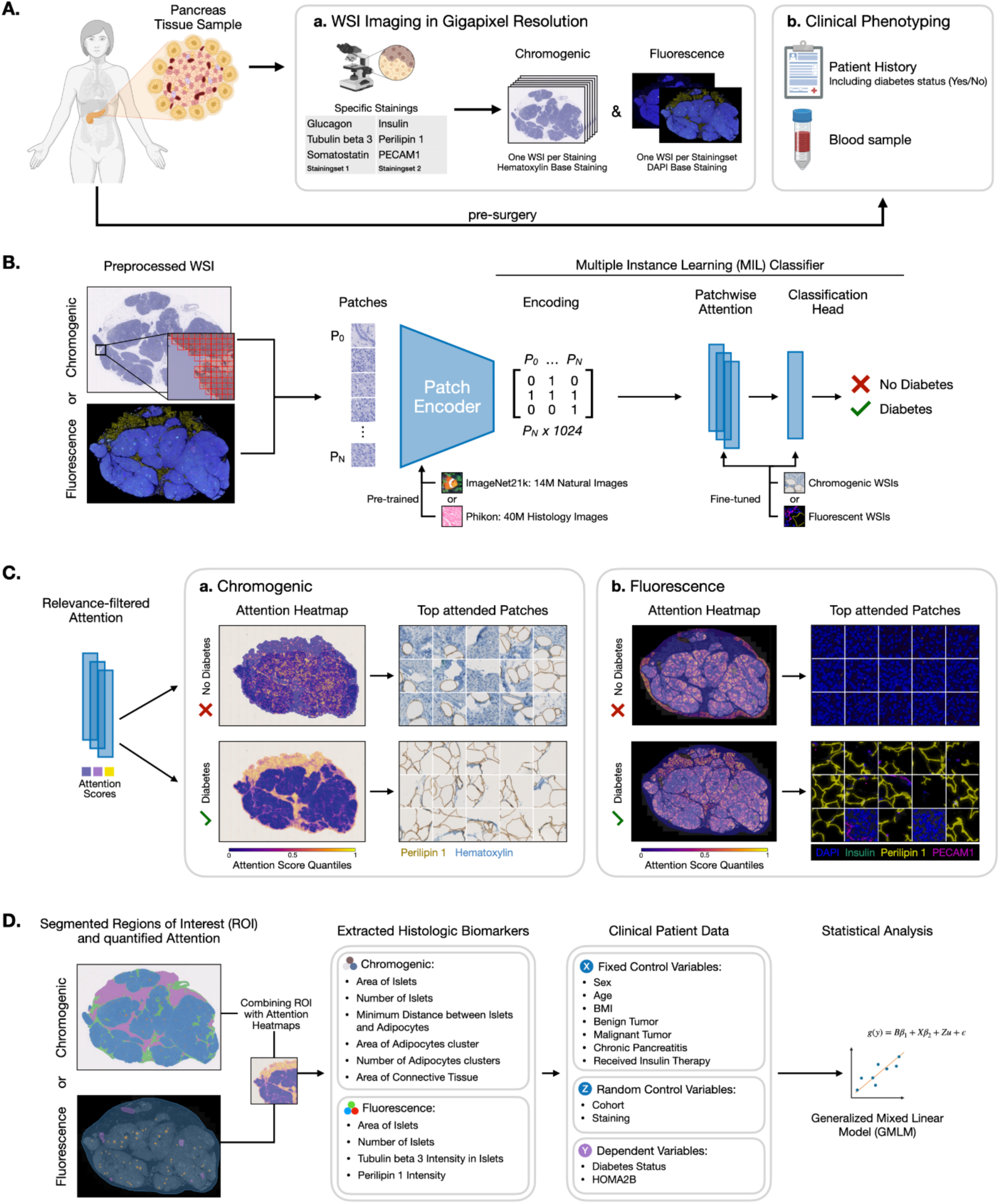
Overview of the analysis procedure from sample and data acquisition to the final statistical analysis. (**A**) Data from immunostained serial pancreatic tissue sections were integrated with fasting blood samples and clinical patient data as described in the methods. Briefly, antibodies against insulin, glucagon, somatostatin, PECAM1, perilipin 1, and tubulin beta 3 were used for single chromogenic immunostaining, whereas in the case of multiplex fluorescence, antibodies were combined into two distinct staining sets. Immunohistological whole slide images (WSIs) were generated via brightfield and fluorescence microscopy. (**B**) Flowchart showing the building process of the models able to predict the T2D status from the WSIs. (**C**) XAI methods were used to identify regions of interest (ROIs) utilized by the models for their prediction. (**D**) The identified ROIs were finally segmented and quantified. The extracted histologic biomarkers, together with covariates from clinical patient data were analyzed with statistical models to assess their association with T2D status and insulin secretion (HOMA2B).

### Classification of diabetes from whole-slide images of pancreatic tissue sections

Subsequent brightfield and fluorescence acquired whole slide images (WSIs) were used to train DL models distinguishing donors with or without T2D (Fig. 1B). Patients were split into a training set (n=75) and a test set (n=25), with the test set used only for the final model evaluation. During model development, we employed a 15-fold cross validation, i.e. 15 different training (n=60 each) and validation (n=15 each) splits to avoid overfitting on certain validation sets. Final test set predictions were obtained by ensembling the 15 individual models, i.e. averaging the individual predictions.

We trained DL models for each of the six chromogenic stainings, while on the fluorescence data, we trained DL models for each of the two stainingsets (Fig. 2). Each of our DL models consisted of two components, the pre-trained feature extractor, and the Multiple Instance Learning (MIL) classifier. Generally, due to their size, each WSI was first divided into smaller-sized squares (patches) and each patch was encoded by the feature extractor. Subsequently, all encodings belonging to one WSI were combined and fed to the MIL classifier which learned to distinguish between T2D and non-T2D based on the combined patch encodings only.

**Figure 2.**
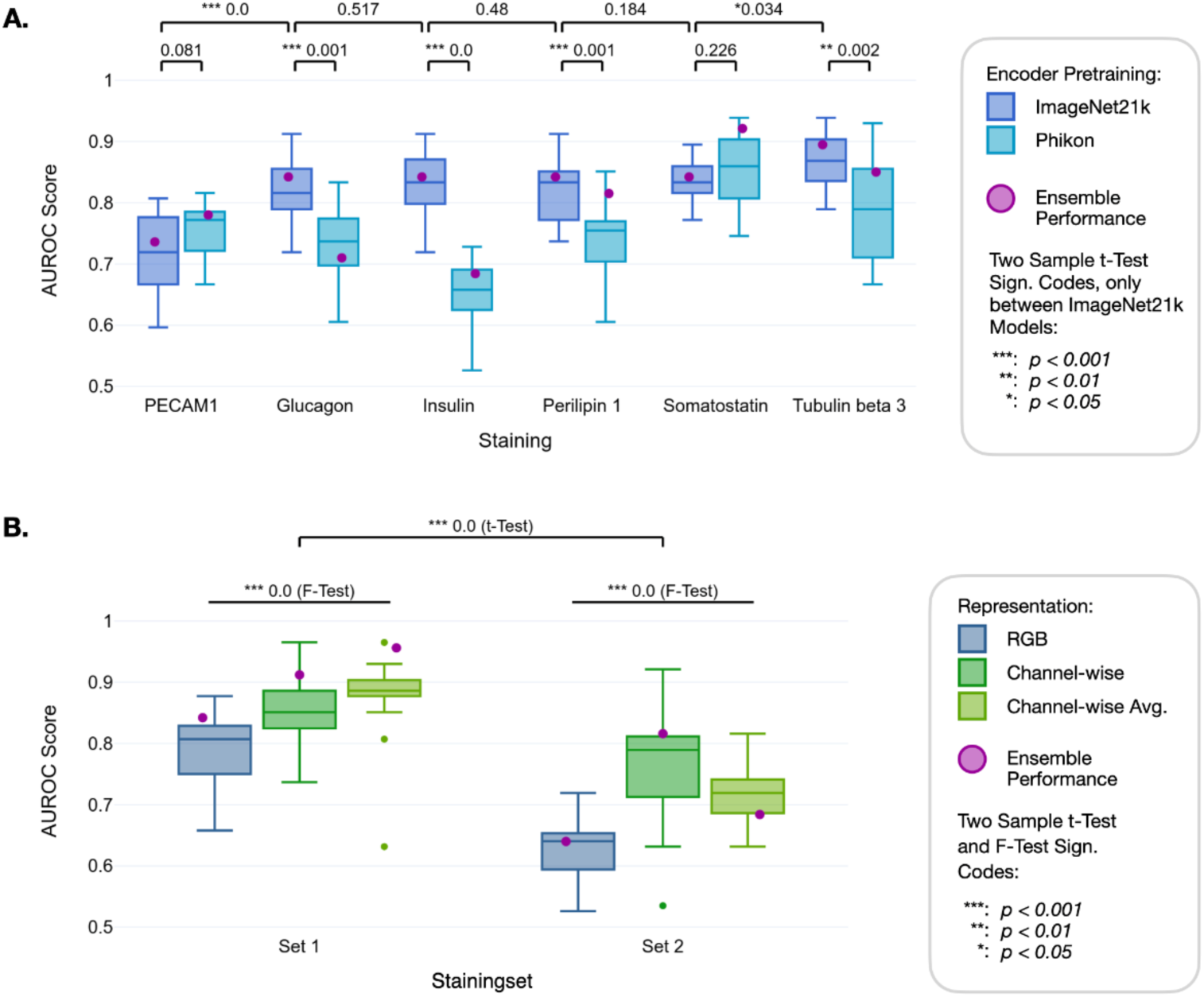
Performance efficiency of the trained models to predict T2D on chromogenic and fluorescence WSIs. Ensemble performance indicates the AUROC achieved when the respective 15 trained models per configuration are combined by aggregating their individual predictions by using the averaged class probabilities to obtain a final prediction. (**A**) AUROCs on the held-out test set (25 patients) across 15 training runs (on 15 folds) showing the performance of ImageNet21k/CLAM with the chromogenic WSIs stained for PECAM1, glucagon, insulin, perilipin 1, somatostatin and tubulin beta 3 compared to Phikon/CLAM. In all cases except PECAM1 and somatostatin ImageNet21k/CLAM yields better performance. Tubulin beta 3 yields the highest performance across stainings (p<0.05), while performance differences between stainings are generally small. (**B**) AUROCs by ImageNet21k/CLAM on fluorescence WSIs stained with Stainingset 1 and 2 and represented in RGB, channel-wise or channel-wise average, showing overall better performance (p<0.001) for Stainingset 1 and better performance (p<0.001) for the channel-wise average representation in that set.

We compared the classification performance for different pre-training approaches on the brightfield WSIs. The ImageNet21k pre-trained (Ridnik et al., 2021) Vision Transformer as feature extractor combined with the CLAM model architecture (Lu et al., 2021) as classifier delivered the best average prediction performance (AUROC = 0.833; Fig. 2A and Suppl. Table 1). Across all stainings except for PECAM1 and somatostatin, there was a significant increase in AUROC when using the ImageNet21k pre-trained encoder. Of note, the chromogenic staining resulting in the best prediction performance was tubulin beta 3 (AUROC = 0.895), while the three islet-typical labels, i.e. insulin, glucagon, and somatostatin showed similar AUROC values (AUROC = 0.842; Fig. 2A, Suppl. Table 1).

On the fluorescence WSIs, we compared three different color-channel representations: “RGB”, “channel-wise” and “channel-wise average” (see Methods section). In short, “RGB” treated the three stainings in a stainingset as rgb color channels processing all channels simultaneously, while the two “channel-wise” representations processed each channel individually and either appended the resulting encodings (“channel-wise”) or averaged them (“channel-wise average”). The latter benefited from co-occurrence information reflecting the spatial relationships between different stainings. When ImageNet21k/CLAM-based prediction was conducted using fluorescence WSIs, Stainingset 1, despite the absence of insulin labeling, yielded significantly better classification results than Stainingset 2. The best diagnostic performance was reached using the “channel-wise average” representation on Stainingset 1 (Ensemble AUROC=0.956; Fig. 2B, Suppl. Table 1).

### AI models attend to specific biological traits

Upon completion of model training, we aimed to understand the biological features utilized by the models in predicting T2D. For this purpose, we employed XAI techniques specifically within the domains of Attention (Chefer et al., 2021) and Attribution (Erion et al., 2020; Ribeiro et al., 2016; Sundararajan et al., 2017) methods, to highlight regions critical to the models when predicting the T2D status (Fig. 1C). We applied these techniques to the best classification model among the evaluated training settings for both fluorescent and chromogenic modalities.

Initially, we focused on determining the significance of specific patches using the built-in attention mechanism of the multiple instance learning (MIL) (Dietterich et al., 1997) classifier and creating attention heatmaps for the individual chromogenic and fluorescence WSIs of each patient (see exemplary Fig. 3). The heatmaps of the chromogenic WSIs revealed both similarities and distinctions in the regions important to the model when classifying a WSI to diabetes or its absence. Specifically, for the “diabetes” outcome (Fig 3A, lower panels), several stainings showed heightened attention to areas abundant in connective tissue (left side of the PECAM1-, glucagon-, insulin-, and somatostatin-stained WSIs) as well as a pronounced focus on adipocytes (top left of the perilipin 1-stained WSI). Both recognized features, i.e. connective tissue enrichment and adipocyte infiltration are mainly localized in the exocrine tissue. Notably, in the chromogenic WSIs the enrichment in connective tissue was revealed by low-intensity diffuse staining of the pancreatic sections by diaminobenzidine precipitates, regardless of antigen specificity, accounting for spatially restricted, high-intensity signals. Conversely, for the “no T2D” outcome, the models devoted significantly less attention to the connective tissue area. Instead, they concentrated on tissue borders and displayed more dispersed attention patterns. The heatmaps for the fluorescence WSIs of both Stainingsets revealed similar attention patterns such as on adipocytes (perilipin 1) in Stainingset 2 (Fig. 3B).

**Figure 3.**
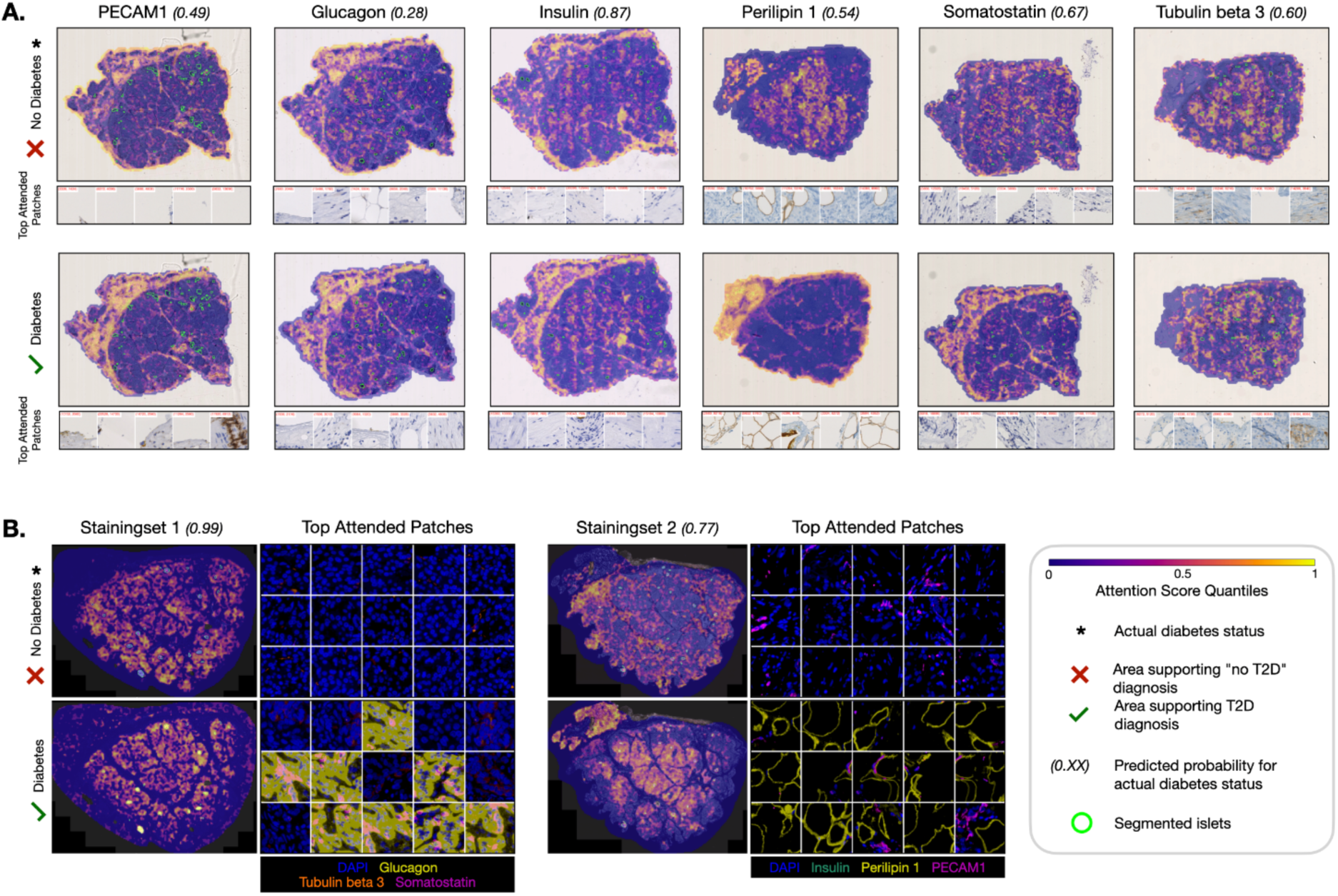
Representative attention heatmaps for the chromogenic and fluorescence WSIs of a single patient. Attention-based heatmaps on global WSI level for (**A**) each chromogenic staining with the respective top 5 attended patches and (**B**) Stainingsets 1 and 2 of fluorescence staining, together with the respective top 15 attended patches associated with T2D or no T2D status.

While these heatmaps facilitate the visualization of larger ROIs in the tissue, they do not provide detailed insights into the finer biological features that the model considers significant. To address this, we first sampled the top attended patches using attention scores (Fig. 3B, Suppl. Fig. 1 for chromogenic WSIs), essentially zooming in from image-level to patch-level. In the case of fluorescence WSIs, these heatmaps reveal that for Stainingset 2, the model indeed exhibits a high level of attention to adipocytes (perilipin 1). However, for Stainingset 1, the model also focuses on islet α-cells (glucagon), when classifying "diabetes" (Fig. 3B). This focus of the model on α-cells when classifying a person as having diabetes is not observable from the patch-level heatmap alone.

To further determine the specific pixel-level features utilized by the model in each of the top-attended patches, we employed multiple attribution methods to quantify the contribution of individual pixels to the predicted outcomes. The model was indeed able to recognize biological traits, such as nuclei, adipocyte cytosolic compartments, inter-cellular structures, or islets (Suppl. Fig. 2). Integrating all XAI findings, we compiled a comprehensive report for each patient, imaging modality, and DL model.

### Quantifying Attention to regions-of-interest

To quantitatively assess the general importance of specific regions across all heatmaps, we implemented a scalable, data-driven methodology. We examined highly attended patches to determine their corresponding tissue structures. Those reoccurring were defined as ROIs and segmented in the WSI using a specifically trained DL segmentation model to enable quantitative analysis of total attention to different relevant tissue features. The attention scores within ROIs were standardized for tissue size and total amount of attention.

Analysis of the attention scores revealed a heterogeneous distribution of attention towards tissue components such as islets, adipocytes, or connective tissue in the individual chromogenic WSIs. Attention to islets showed the highest z-scores for insulin- and glucagon-stained WSIs, independent of diabetes status (Fig. 4A). The attention to adipocytes showed significant variability across different stainings, with the highest z-scores for the adipocytes specific perilipin 1 staining (Fig. 4B). Notably, the model’s attention to connective tissue in the chromogenic WSIs was significantly higher in patients with T2D on the PECAM1, perilipin 1 and tubulin beta 3 stainings, indicating its potential relevance in the pathology of the disease (Fig. 4C).

**Figure 4.**
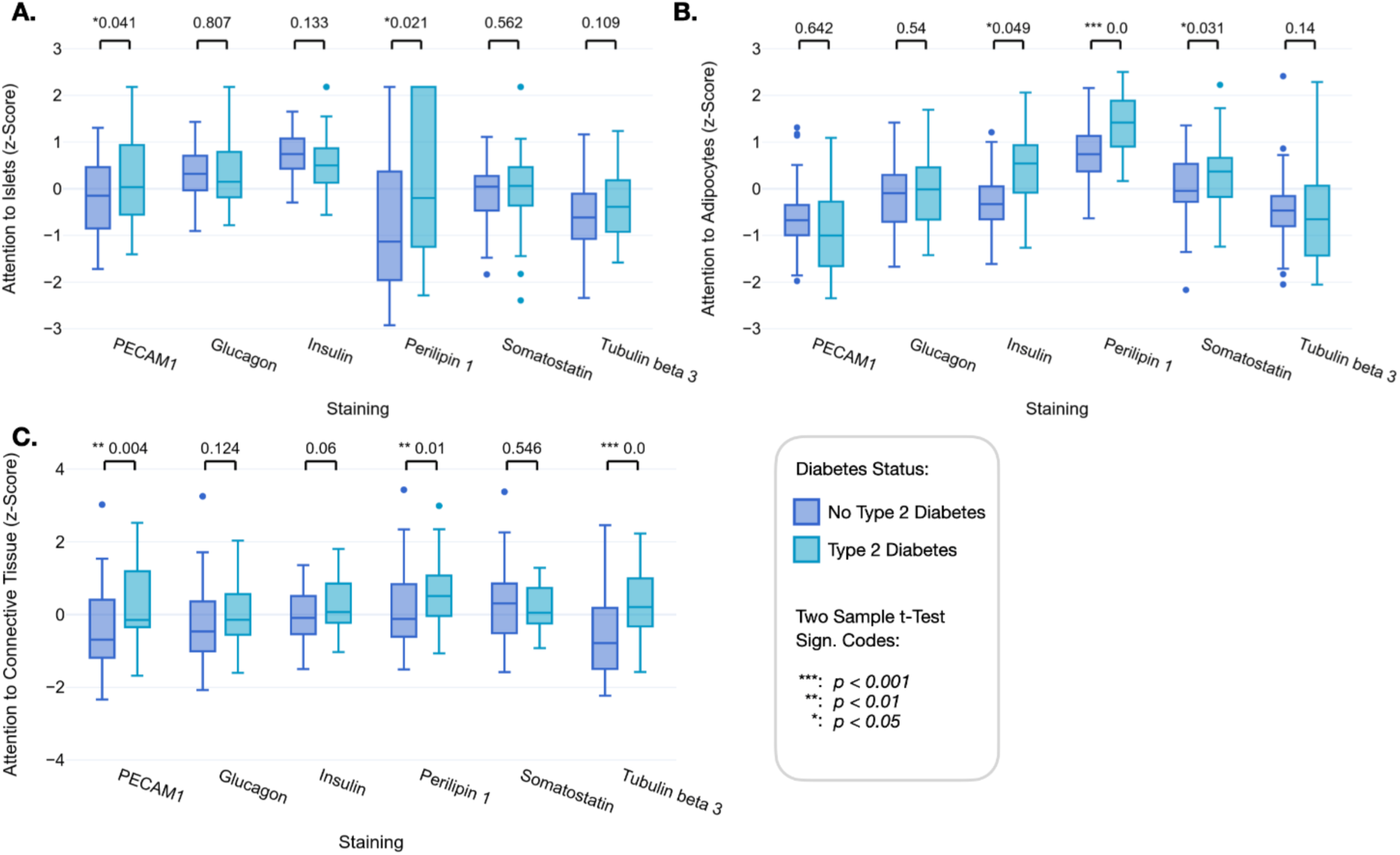
Quantifying attention to regions of interest for chromogenic WSIs of non-diabetic and T2D donors. Z-scores for average attributed attention to (**A**) islets, (**B**) adipocytes, and (**C**) connective tissue-rich areas for the individual chromogenic stainings of patients with and without T2D. Respective p-values (T-test) indicate significant differences between T2D and non-diabetes status. Variables on non-interpretable Y scales are z-standardized.

In the fluorescence WSIs, total attention and attention to islets differed only between the stainings of Stainingset 2, while the models attributed significantly higher attention to tubulin beta 3, perilipin 1 and glucagon in the WSIs of T2D donors (Fig. 5A). The ratio between attention to islets and total attention to all patches containing stained tissue (which are only the patches containing islets in the case of glucagon, insulin and somatostatin) shows that islets are attended above average for tubulin beta 3 and PECAM1 and below average for perilipin 1 staining compared to the rest of the tissue (Fig. 5C).

**Figure 5.**
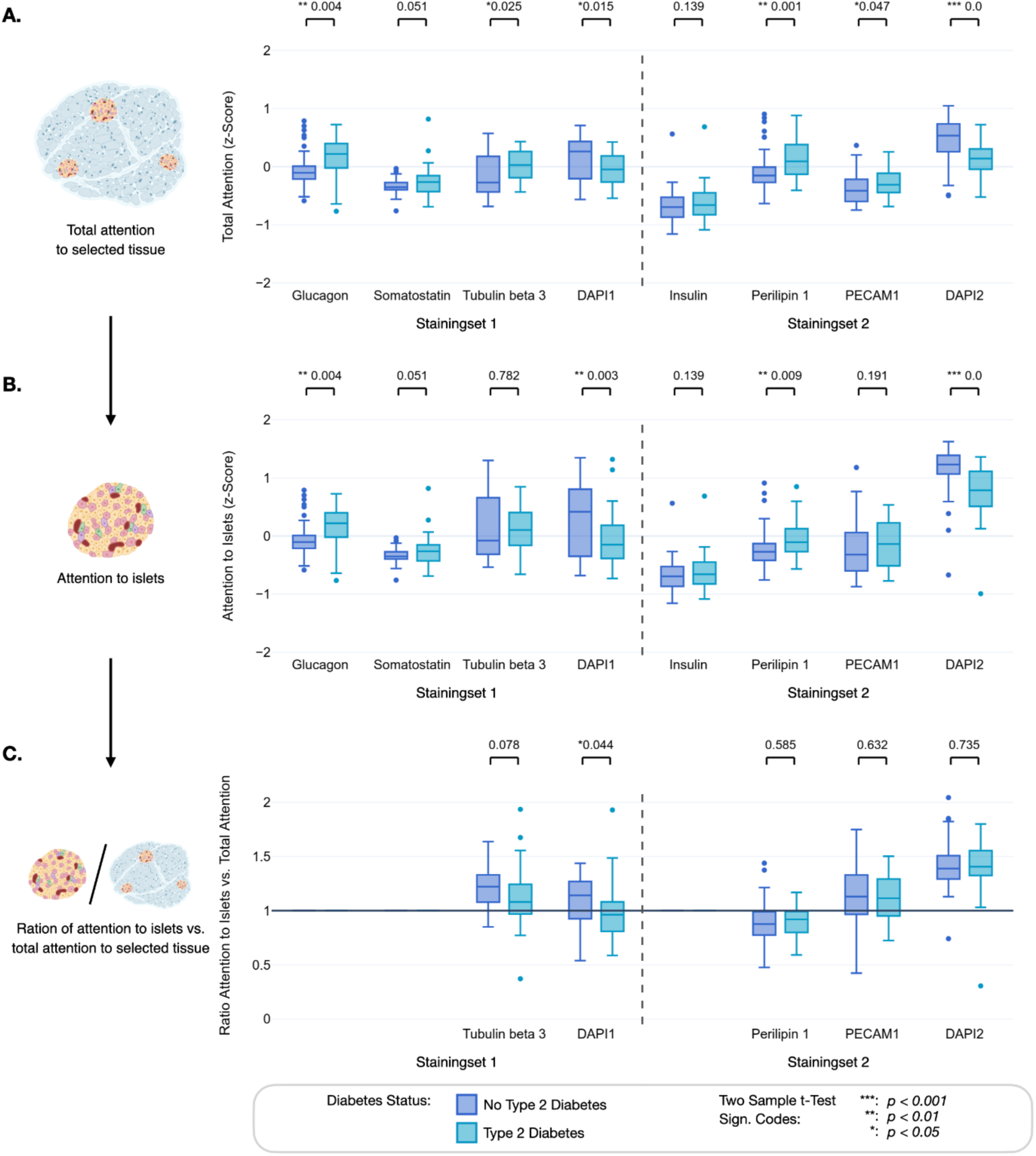
Comparison of attention within the total area containing stained tissue and islets between the individual fluorescence stainings. (**A**) Z-score for the attributed attention of the channel-wise models to the different staining channels in the fluorescence WSIs labeled with Stainingsets 1 and 2. (**B**) Attention of the channel-wise models to islets in the fluorescence WSIs. (**C**) Ratio of attention to islets and the total attention to the total area containing stained tissue. Values greater than 1 suggest that, for this staining, the islets are more important to the model than the average tissue patch. The ratio is always 1 for stainings that exclusively stain the islets, i.e. glucagon, somatostatin, and insulin. Respective p-values (T-test) indicate significant differences between T2D and non-diabetes status. Variables on non-interpretable Y scales are z-standardized.

### Regions-of-interest and XAI-derived results provide quantifiable histologic biomarkers

Based on the assessment of the XAI results, we define various histologic biomarkers for the chromogenic and fluorescence modalities, which we compute through the quantitative XAI results and the segmentation maps (Fig 1D). Notably, these biomarkers exhibited variations between both modalities as certain ROIs could not be computed for fluorescent WSIs, e.g., connective tissue due to the absence of a dedicated staining. All histologic biomarkers were standardized for tissue size.

A first explorative analysis of the histologic biomarkers revealed that persons without diabetes generally exhibited larger islets (Fig. 6A). In the case of perilipin 1, the segmentation of the islets was more challenging due to a lack of islet-specific staining, resulting in considerably fewer detected islets. Further, patients with T2D tended to have larger adipocyte clusters (Fig. 6B) and connective tissue-enriched areas (Fig. 6C). An adipocyte cluster was defined as either connected accumulations of adipocyte cells or a single non-connected adipocyte cell in the tissue. When assessing the average minimum distance between each islet and the nearest adipocyte, computed only on the insulin staining segmentations, we observed that in patients with T2D, the islets were significantly closer to adipocytes than in patients without T2D (Fig. 6D).

**Figure 6.**
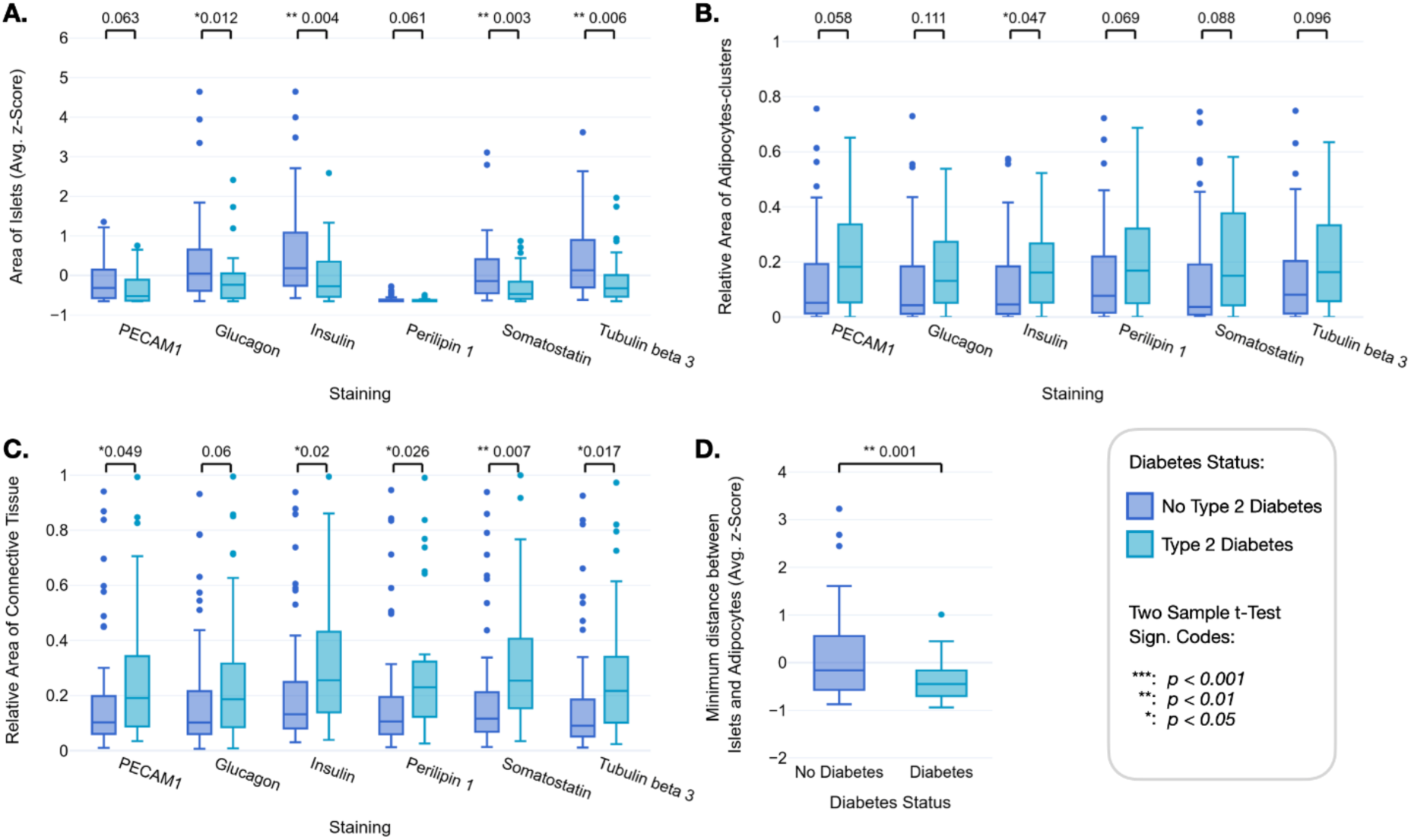
Quantification of XAI-derived histologic biomarkers for chromogenic WSIs of non-diabetic and T2D donors. (**A**) Average islet area, and relative areas of (**B**) pancreatic adipocyte clusters (a cluster was defined as connected adipocyte cells, single non-connected adipocyte cells counted as one cluster), and (**C**) connective tissue in the chromogenic WSIs of patients with and without T2D. (**D**) The average minimum distance between each islet and the nearest adipocyte in the insulin-stained WSIs from non-diabetic and T2D donors. Respective p-values (T-test) indicate significant differences between T2D and no T2D status in each plot.

### Statistical analysis reveals a significant association of histologic biomarkers with diabetes status and insulin secretion

To statistically assess the impact of the histologic biomarkers on diabetes status and insulin secretion, measured through HOMA2B, we considered several control variables, which were either directly pertaining to individual donor traits, including sex, age, body mass index (BMI), underlying pancreatic tumor, chronic pancreatitis, and insulin therapy; or experimental conditions, including cohort, and tissue staining (Fig. 1D). We analyzed the data using generalized mixed linear models (GMLMs), accounting for random effects linked to the cohort and staining method.

For the chromogenic stainings, regression analysis revealed a significant positive association of both adipocyte cluster areas (*0.266; p=0.035*) and connective tissue-rich areas (*0.524; p<0.001*) with diabetes status (Table 1A). We observed an inverse association of diabetes status with the number of adipocyte clusters (*-0.401; p=0.011*). The distance between islets and adipocytes also showed an inverse association (*-0.633; p=0.007*), suggesting that a smaller distance between adipocytes and islets is associated with T2D. For the fixed effects control variables, sex (males vs females) (*0.610; p=0.007*), age (*0.741; p<0.001*), BMI (*0.538; p<0.001*), and chronic pancreatitis (*1.420; p<0.001*) had a significant association with diabetes status.

**Table 1:**
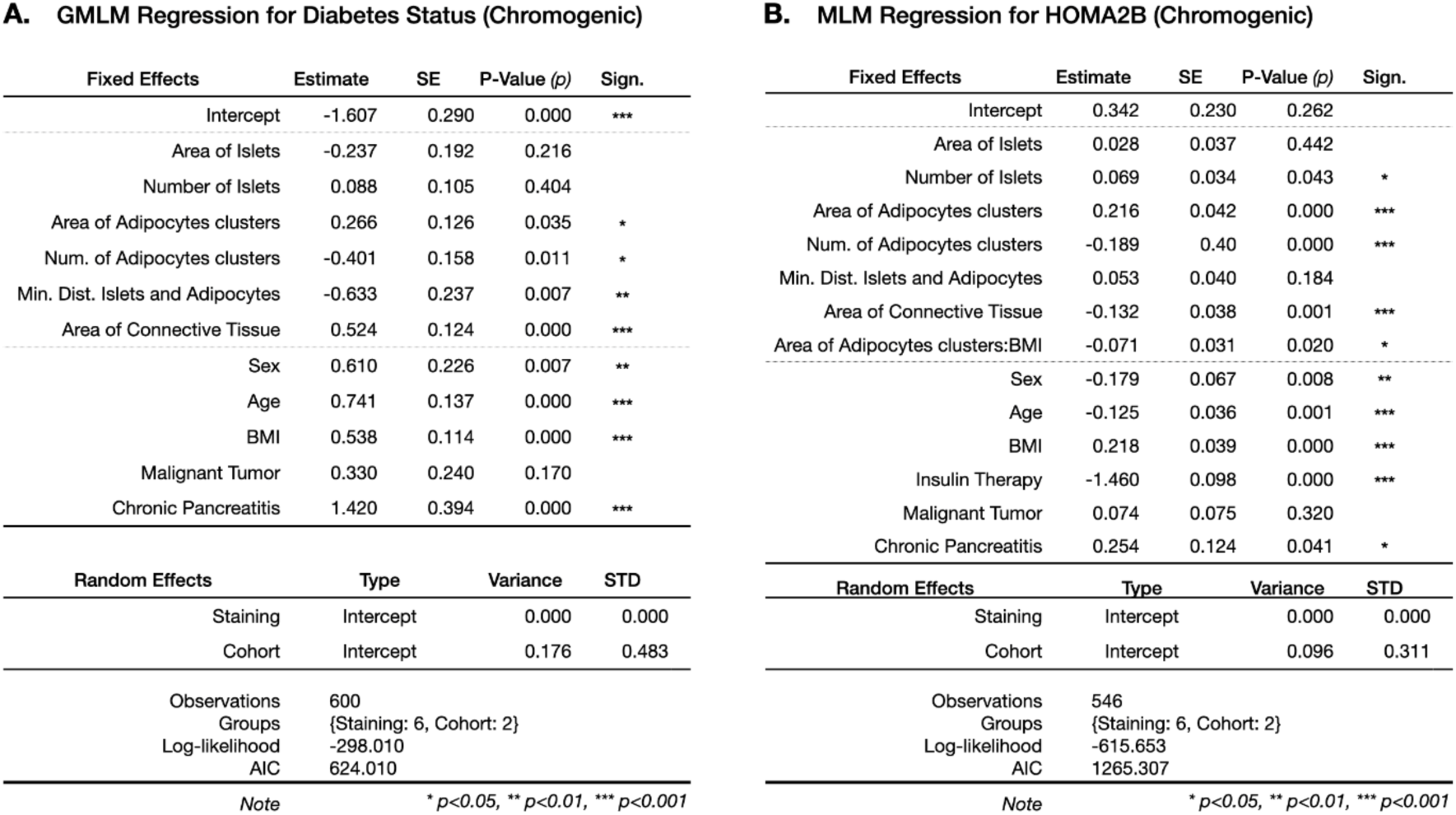
Regression results of the generalized mixed linear model analysis for (**A**) diabetes status and (**B**) HOMA2B levels based on chromogenic stainings. All continuous variables were standardized, with a mean of 0 and a standard deviation of 1, to ensure comparability across measures.

**A. GLMM Regression for Diabetes Status (Chromogenic)**
| Fixed Effects | Estimate | SE | P-Value (p) | Sign. |
| --- | --- | --- | --- | --- |
| Intercept | -1.607 | 0.290 | 0.000 | *** |
| Area of Islets | -0.237 | 0.192 | 0.216 |  |
| Number of Islets | 0.088 | 0.105 | 0.404 |  |
| Area of Adipocytes clusters | 0.266 | 0.126 | 0.035 | * |
| Num. of Adipocytes clusters | -0.401 | 0.158 | 0.011 | * |
| Min. Dist. Islets and Adipocytes | -0.633 | 0.237 | 0.007 | ** |
| Area of Connective Tissue | 0.524 | 0.124 | 0.000 | *** |
| Sex | 0.610 | 0.226 | 0.007 | ** |
| Age | 0.741 | 0.137 | 0.000 | *** |
| BMI | 0.538 | 0.114 | 0.000 | *** |
| Malignant Tumor | 0.330 | 0.240 | 0.170 |  |
| Chronic Pancreatitis | 1.420 | 0.394 | 0.000 | *** |
| Random Effects | Type | Variance | STD |  |
| Staining | Intercept | 0.000 | 0.000 |  |
| Cohort | Intercept | 0.176 | 0.483 |  |
| Observations | 600 |  |  |  |
| Groups | {Staining: 6, Cohort: 2} |  |  |  |
| Log-likelihood | -298.010 |  |  |  |
| AIC | 624.010 |  |  |  |
| Note | | * $p < 0.05$, ** $p < 0.01$, *** $p < 0.001$ | | |

| Fixed Effects | Estimate | SE | P-Value (p) | Sign. |
| --- | --- | --- | --- | --- |
| Intercept | 0.342 | 0.230 | 0.262 |  |
| Area of Islets | 0.028 | 0.037 | 0.442 |  |
| Number of Islets | 0.069 | 0.034 | 0.043 | * |
| Area of Adipocytes clusters | 0.216 | 0.042 | 0.000 | *** |
| Num. of Adipocytes clusters | -0.189 | 0.40 | 0.000 | *** |
| Min. Dist. Islets and Adipocytes | 0.053 | 0.040 | 0.184 |  |
| Area of Connective Tissue | -0.132 | 0.038 | 0.001 | *** |
| Area of Adipocytes clusters:BMI | -0.071 | 0.031 | 0.020 | * |
| Sex | -0.179 | 0.067 | 0.008 | ** |
| Age | -0.125 | 0.036 | 0.001 | *** |
| BMI | 0.218 | 0.039 | 0.000 | *** |
| Insulin Therapy | -1.460 | 0.098 | 0.000 | *** |
| Malignant Tumor | 0.074 | 0.075 | 0.320 |  |
| Chronic Pancreatitis | 0.254 | 0.124 | 0.041 | * |
| Random Effects | Type | Variance | STD |  |
| Staining | Intercept | 0.000 | 0.000 |  |
| Cohort | Intercept | 0.096 | 0.311 |  |
| Observations | 546 |  |  |  |
| Groups | {Staining: 6, Cohort: 2} |  |  |  |
| Log-likelihood | -615.653 |  |  |  |
| AIC | 1265.307 |  |  |  |
| Note | | * $p < 0.05$, ** $p < 0.01$, *** $p < 0.001$ | | |

When regressing on insulin secretion, we observed a significant negative association with the area of connective tissue (*-0.132; p=0.001*) and the number of adipocyte clusters (*-0.189; p<0.001*) (Table 1B). Contrarily, there was a significant positive association of insulin secretion with the number of islets (*0.069; p=0.043*) and the area of adipocyte clusters (0.216, p<0.001). This association turns negative when interacting with BMI, suggesting an attenuation of this association in the case of higher BMI (*-0.071; p=0.020*). Among the control variables, sex (*-0.179; p=0.008*), age (*-0.125; p=0.001*), BMI (*0.218; p<0.001*), insulin therapy (*-1.460; p<0.001*), and chronic pancreatitis (*0.254; p=0.041*) had a significant association with insulin secretion. Among the random effects in both models, there were large variances between cohort intercepts and virtually no variances across different stainings.

Results from fluorescence WSIs are only partially comparable to those from chromogenic data. One reason is the inability to detect connective tissue-rich structures in the absence of specific fluorescent staining. In the case of the chromogenic stained WSI, we compute each biomarker for every different staining, resulting in six sets of imaging markers per patient. However, the two multiplexed WSIs resulted in only one set of imaging markers per patient, as the biomarkers of the two stainingsets were distinct except for the area or size of the islets which was computed over both sets, reducing the sensitivity of the fluorescence statistical models. When regressing the fluorescence biomarkers on diabetes status, only the control variables age (*0.832; p=0.012*), BMI (*0.593; p=0.024*) and chronic pancreatitis (*1.915; p=0.048*) were significant (Suppl. Table 3A). However, when analyzing insulin secretion, we observed a significant association with the area of islets (*-0.207; p=0.030*) and the number of islets (*0.280; p=0.008*), as well as the control variables BMI (*0.243; p=0.008*) and insulin therapy (*-1.435; p<0.001*) (Suppl. Table 3B). We refer to Suppl. Fig. 7 and 8 for the results of all GMLMs.

## Discussion

Our study aimed to uncover biological insights into T2D-associated changes in human pancreas tissues combining diverse immunostainings and AI techniques. To this end, we trained extensive DL models to differentiate between the presence or absence of T2D from gigapixel-sized histopathologic WSIs of pancreatic tissues from living donors recruited at two academic centers. This process involved multiple immunohistological stainings combined with two different microscopy techniques. We subsequently employed XAI methods to find AI-attended ROIs from these WSIs, from which we computed and analyzed histologic biomarkers using exploratory visualization and GMLMs.

The AI-based T2D prediction models demonstrated a highly reliable performance, depending on the staining approach. The highest predictive performance was obtained with multiplex fluorescent staining using our predefined Stainingset 1, comprising stainings for ɑ- and ẟ-cells, i.e., for glucagon and somatostatin, respectively, as well as staining of tubulin beta 3 for neuronal-axonal structures. The predictive power of this Stainingset was surprisingly better than that of Stainingset 2 which specifically marked insulin, i.e. β-cells, in addition to adipocyte lipid droplet membranes and endothelial cells via perilipin 1 and PECAM1 staining, respectively. We also saw differences between the techniques aggregating information from multiplex stainings. While the ‘channel-wise’ approach did not retain co-occurrence information of the different stainings, the ‘channel-wise average’ approach reintroduced this information by using mean encodings for each pixel location of the WSI at the trade-off of losing channel-specific details. The clear benefit of the ‘channel-wise average’ approach in the case of Stainingset 1 likely resulted from its provision of more robust information on islets compared to Stainingset 2 due to the co-occurrence of two different channels (glucagon, somatostatin), highlighting islets only, as well as tubulin beta 3. Of note, tubulin beta 3 is not exclusively restricted to neuronal axons, but is also present, albeit at a much lower level, in islet cells, including β-cells (Suppl. Fig. 8) (Xin et al., 2016). Thus, Stainingset 1 may also have indirectly incorporated information on non-ɑ, non-ẟ islet cells, including β-cells, and thereby, identify subtle changes in the relationship among these cells and in the islet structure. On the other hand, Stainingset 2 had an inferior prediction performance with the channel-wise averaging approach compared to the channel-wise approach because it employed three stainings mostly occurring in non-overlapping areas, i.e. β-cells, adipocytes, and endothelial cells and thus, its benefit from co-occurrence information was lower.

While the chromogenic stainings had lower predictive performances compared to the immunofluorescence images, their highest average predictive performance was found for tubulin beta 3, which was also a component of the best-performing fluorescence Stainingset 1. Of note, when comparing the average attention to each of the fluorescent stainings between samples without and with T2D, we saw higher attention to tubulin beta 3 in T2D compared to non-diabetic WSIs (Fig. 5A). The data suggest that alterations in tubulin beta 3 stained structures could constitute a distinguishing pattern of T2D as islet innervation has an impact on islet cell function (Alvarsson et al., 2020; Hampton et al., 2022). Indeed, through the autonomic nervous system, brain-derived signalsreach pancreatic islet cells, including β-cell primary cilia (Alvarsson et al., 2020; Hampton et al., 2022), and modulate insulin secretion in humans (Heni, 2024). Our current data suggest that alterations in these pathways at the level of innervation of the islet might be involved in T2D.

The islets received variable levels of attention within the individual chromogenic and multiplexed fluorescence stainings. While in the chromogenic analysis islets stained for either insulin or glucagon received the greatest attention (Fig. 4A), in the case of the fluorescence Stainingset 1, the glucagon signal received the highest attention relative to somatostatin and tubulin beta 3 (Fig. 5B). The ratio between the attention to the islets (Fig. 5B) and the total attention to the selected tissue patches (Fig. 5A), which also includes exocrine tissue, indicated that among non-islet restricted markers, tubulin beta 3 and PECAM1 were attended above average (Fig. 5C). These findings are consistent with the notion that islets are primarily altered in T2D relative to the surrounding pancreatic tissue. For PECAM1, this likely reflects changes in vessel cytoarchitecture, but for tubulin beta 3, it’s unclear whether the observed alterations stem from neuronal fibers, islet cells, or both. With the chromogenic stainings for insulin, glucagon, somatostatin, and tubulin beta 3 we observed significantly smaller islets in patients with T2D than without (Fig. 6A). Although we cannot say whether the reduction of β- or ɑ-cell mass accounts for the smaller islets, previous observations suggested that larger islets are preferentially lost in T2D (Kilimnik et al., 2011). Intriguingly, some evidence indicates that smaller islets have better glucose-responsive insulin secretion compared to large islets (Steffen et al., 2011; Fujita et al., 2011; Farhat et al., 2013). This is in line with our finding that lower insulin secretion was associated with larger fluorescence image-derived islet size when also adjusting for confounders such as number of islets, age, BMI, underlying disease, and exogenous insulin therapy.

Attention differences in perilipin 1 staining, along with combined statistical analysis of factors contributing to T2D highlight intra-pancreatic adipocytes as key tissue compartments related to the disease. In our study, we used two histologic biomarkers of intra-pancreatic steatosis: the area of adipocyte clusters and the number of adipocyte clusters. The correlation coefficient between these biomarkers is only 0.4, suggesting that they reflect different aspects of intra-pancreatic steatosis. The analysis of the chromogenic biomarkers revealed that the area of adipocyte clusters is associated with T2D (Table 1A, Fig. 6B), while the number of adipocyte clusters was inversely correlated with the condition (Table 1A). The area of adipocyte clusters was well correlated with BMI, whereas there was no substantial correlation between the number of adipocytes and BMI (Suppl. Fig. 12). For insulin secretion, the association of these biomarkers suggests higher insulin secretion with a larger area of adipocyte clusters and lower insulin secretion with a higher number of adipocyte clusters (Table 1B). Of note, insulin hypersecretion to compensate for insulin resistance is an early feature of T2D, and a positive association of the area of intra-pancreatic adipocytes with insulin secretion was observed in persons with a low genetic risk of T2D (Wagner et al., 2020). A higher number of adipocyte clusters may indicate a more advanced adipose infiltration of pancreatic tissue, i.e. intralobular adipocytes (Wright et al., 2023). Such an intralobular infiltration would allow the adipocytes to get closer to the islets and thereby alter the islet microenvironment, with negative consequences for insulin secretion (Suppl. Fig. 3). Indeed we observed that in patients with T2D, pancreatic islets are on average closer to adipocytes than in patients without T2D, i.e. the distance between islets and adipocytes is negatively correlated with the T2D status (Fig. 6D, Table 1). These associations argue for paracrine effects of the adipocytes secretome impacting islets and contributing to T2D (Gerst et al., 2019).

Intra-pancreatic steatosis has been known to be associated with T2D risk (Wagner et al., 2021). However, this association is substantially modulated by factors such as genetic risk or prevailing metabolic background (Heni et al., 2010; Wagner et al., 2020). Our results also unveiled an interaction between the area of adipocyte clusters and BMI, suggesting a diminishing effect of intra-pancreatic steatosis on insulin secretion in more obese persons (Table 1B). This effect modification of BMI on intra-pancreatic steatosis is consistent with epidemiologic data from a Japanese cohort showing that pancreatic fat robustly predicts diabetes only in lean individuals (Yamazaki et al., 2020).

Although we did not use a specific collagen staining, the peroxidase reaction, employed for antigen visualization in the chromogenic approach, enabled co-visualization of the surrounding reference space including fibers in ECM (extracellular matrix) -enriched areas within the pancreas tissue, as previously reported for other tissue types (de Haan et al., 2021). Combined with hematoxylin staining detecting cell nuclei, this led to a clear distinction between the lobular acinar tissue and the intercalated connective tissue. In the WSIs labeled with islet cell non-specific stainings, i.e. PECAM1 and perilipin 1, the DL models showed greater attention to these fibrotic-like, connective tissue-enriched areas when predicting T2D versus the no T2D state (Fig. 4C). Furthermore, our histologic biomarkers analysis revealed more abundant connective tissue-enriched areas, a fibrotic hallmark, in WSIs of T2D (Fig. 6C), in accordance with previous findings. Fibrosis has indeed been recognized as a structural alteration in the pancreas of persons with T2D (Deng et al., 2004; Hayden, 2007; Shimizu et al., 2019; Wright et al., 2023) and ECM-related genes are upregulated in islets of living donors with T2D (Wigger et al., 2021).

When assessing the relationship of the histologic biomarkers with T2D and insulin secretion, we found a positive association of fibrotic area with T2D and lower insulin secretion as measured by HOMA2B (Table 1). Of note, these associations were independent of the underlying pancreatic tumor, whether benign or malignant, chronic pancreatitis, and insulin therapy. According to our results, pancreatic tissue of patients with T2D undergo morphological alterations that extend beyond islets, such as enrichment in fibrotic-like structures and more islet-proximal adipocytes, which most likely have negative consequences for the islet microenvironment and islet function.

While this data-driven approach has the potential to accelerate and facilitate the understanding of T2D pathogenesis, several potential limitations must be considered. One is that immunostaining approaches can be associated with technical pitfalls such as antibody binding specificity, nonspecific background staining, or variations in the staining intensity across samples regardless of target antigen level. Furthermore, our histological preparations were not representative of the whole organ, since they were obtained from limited regions of the pancreas, the anatomical location of which varied among the donors. Further, despite originating from the “healthy” margins of the surgical resection, as also verified by postoperative pathological assessment, the samples could be affected by changes in the tissue microenvironment such as inflammation, fibrosis, and angiopathy (Korpela et al., 2022).

In our study, we analyzed a unique WSI dataset of the human pancreas, derived from two independent cohort sites, encompassing pre-surgical metabolic phenotyping of living donors both with and without T2D. Our comprehensive staining approach covered a wide spectrum of pancreatic morphology, including islets, innervation, adipocytes, and vascularization, captured with brightfield and multiplex-fluorescence microscopy. Based on this unique dataset, we trained several DL models that, unlike a routine pathological assessment, reliably allow the prediction of the T2D status. This reliable prediction is already novel and represents a remarkable advancement in the field of DL-based histologic biomarker discovery, especially because the pancreas, although a central organ in the pathophysiology of T2D, only exhibits subtle anatomical changes, unlike in the case of cancer or pancreatitis. Further, our quantitative analysis uncovered histologic biomarkers that support existing hypotheses about the involvement of pancreatic islets and their innnervation, adipocytes, and enrichment of connective tissue in T2D. Although β-cells are considered to play the key role in T2D, α- and δ-cells have gained increased attention during the last years, being essential intra-islet modulators of β-cell function (Campbell et al., 2020; Marchetti et al., 2012). Remarkably, the most accurate prediction was obtained with a stainingset that focused on islet α- and δ-cells and neuronal axons only, without specific attention to β-cells. The findings highlight the complexity of T2D pathology and shall further motivate the deployment of XAI methods on other cohorts in the unbiased identification of novel structural histologic biomarkers. This could guide our focus in the research of diabetes prevention and treatment on the most promising targets.

## Methods

### Clinical Cohort

We analyzed clinical, laboratory, and histologic data obtained within the “Studying Islets from Living Donors” (SILDS) programs at two academic sites of the German Center for Diabetes Research network, the University Hospital Tübingen and the University Hospital Dresden (Suppl. Fig. 4 and 5). Patients undergoing pancreatic surgery for different indications provided written informed consent to donate blood samples and pancreas tissue, and share health records and laboratory data for research purposes at both study sites (Tübingen and Dresden). The study was approved by the Ethical Committees of the Technische Universität Dresden (Reference EK 151062008) and Eberhard Karls Universität Tübingen (Reference 697/2011BO1). We obtained macroscopically healthy tissue resected during surgery but not required for further pathology workup. All patients were of European ethnicity. Additionally, fasting blood was drawn pre-surgery for detailed metabolic phenotyping. Fasting glucose and C-peptide levels were measured as previously described (Babbar et al., 2018), and homeostatic model assessment (HOMA) of insulin secretion was calculated using the computer model-based HOMA-2B using glucose and C-peptide (Levy et al., 1998). None of the participants had depleted endogenous insulin production as measured by C-peptide-based HOMA2B (lowest HOMA2B: 6 with a diabetes duration of 24 years), excluding type 1 diabetes among the participants. Information on medical history was collected by a physician. Documented by their health records, T2D patients were diagnosed as having T2D at least one year before admission to pancreatic surgery. This excludes diabetes in the context of exocrine pancreatic disease. In contrast, patients without diabetes neither had diabetes nor did they fulfill diagnostic criteria of T2D based on glycated hemoglobin (HbA1c) and fasting glucose, as defined by the American Diabetes Association (American Diabetes Association, 2020).

### Data Acquisition

#### Hospital Patient Data

The patient cohort-related metadata are summarized in Suppl. Fig. 6 and Suppl. Table 2. The quantitative analysis of these parameters in Suppl. Fig. 6 revealed an unbalanced distribution of sex, age, BMI, tumor type, chronic pancreatitis, and cohort between the patients with and without diabetes. In 9 patients, fasting blood samples were not obtained prior to surgery and therefore HOMA2B was not calculated.

#### Chromogenic immunostaining and brightfield microscopy

Formalin-fixed, paraffin-embedded (FFPE) pancreatic sections (2-4 µm thick) were processed using an automatic slide stainer BenchmarkUltra (Ventana Technology, Roche Diagnostics). Deparaffinization was performed for four min at 72°C using EZPrep (Roche Ventana, #5279771001), followed by antigen-retrieval (AR) for 40 minutes at 100°C with TRIS-based CC1-buffer (Roche Ventana, #5424569001). After peroxidase-inhibition with I-View Inhibitor (Roche, #06396500001), the sections were incubated with primary antibodies against insulin (1:1000; Dako, #A0564), glucagon (1:600; Santa Cruz, #sc13091), somatostatin (1:6000; Invitrogen, #14-9751-80), CD31 (1:100; Dako, #M0823), perilipin 1 (1:2000; Progen, #651156), and tubulin beta 3 (1:2500; R&D Systems, #MAB1195). The secondary horseradish peroxidase-linked antibody was detected via an Opti-View DAB IHC detection kit (Roche Ventana, #06396500001). The samples were counterstained with hematoxylin. WSI acquisition was performed with a Hamamatsu NanoZoomer 2.0-HT using 20x magnification and NDP.scan software. Exemplary zoom-ins of WSIs are shown in Suppl. Fig. 7.

#### Multiplex fluorescent staining and fluorescence microscopy

Consecutive 2-4 µm FFPE sections derived from the identical pancreatic specimens used for the chromogenic stainings underwent fluorescent staining via an automated system DiscoveryUltra (Ventana Technology, Roche Diagnostics). Deparaffinization utilized EZPrep (Roche Ventana; #5279771001) for 32 min at 69°C, and subsequent AR was performed with TRIS-based CC1-buffer (Roche Ventana, #5424569001) at 91°C for 48 min. Primary antibody cocktails were applied after incubation with a human FC-receptor-blocking reagent (1:50; Miltenyi, #130-059-901).

The antibody cocktails for Stainingset 1 included for the first incubation antibodies against glucagon (1:200; Abcam plc. #Ab10988) and tubulin beta 3 (1:25; R&D Systems, MAB1195), for the second incubation mouse IgG1 con. AF555 (1:200; Invitrogen, #A-21127) and mouse IgG2a con. AF647 (1:50; Invitrogen, #A21241), and for the third incubation somatostatin con. AF750 (1:200; Novus Biologicals, #NBP2-99309 AF750) with DAPI (1 ng/ml; Invitrogen, #D1306). The cocktails for Stainingset 2 included for the first incubation antibodies against perilipin 1/PLIN1 (1:50; Progen, #690156) and PECAM1/CD31 (1:33; Abcam, #ab134168), for the second incubation mouse IgG1 con. AF555 (1:100; Invitrogen, #A21127) and rabbit IgG con. AF750 (1:50; Invitrogen, #A21039), and for the third incubation insulin con. AF488 (1:200; Invitrogen, #53-9769-82) with DAPI (1 ng/ml; Invitrogen, #D1306). WSI acquisition was performed with a slide scanner (Zeiss AxioScan.Z1 equipped with ZEN 3.10 software of the Light Microscopy Facility, a Core Facility of the CMCB Technology Platform at TU Dresden.) at 20x magnification. Exemplary WSIs are shown in Suppl. Fig. 8.

### Deep Learning Models

Model Architectures. Training DL models on WSIs requires special precautions due to the large file size. Since an entire image does not fit into GPU (graphics processing unit) memory, a patch-based approach is required, where patches are similar-sized crops of pixel values of tissue regions in the WSI. A difficulty to this approach is that only a global label per image is available but not per patch. It is not known if the manifestation of the respective image-level class is actually given in every patch or if there are also patches that are not indicative of the image-level class and would hence confuse the models during training. A Multiple Instance Learning Approach (MIL) is commonly used to tackle such problems (Dietterich et al., 1997). In MIL the individual patches are first encoded by a pre-trained feature extractor and subsequently pooled and fed together to a MIL classification algorithm that is trained to predict the image-level class label. We tested two different feature extractors on the chromogenic WSIs. The first is a Vision Transformer pre-trained on ImageNet21k (Ridnik et al., 2021), which consists of natural images, while the second is Phikon (Filiot et al., 2023), a Vision Transformer pre-trained on over 6 thousand histologic WSIs and therefore specifically tailored to the domain. As the MIL algorithm, we used Clustering-constrained Attention Multiple Instance Learning (CLAM) (Lu et al., 2021) as well as Chowder (Courtiol et al., 2020). Both architectures deal with the variability in patch importance in different ways. CLAM uses an attention mechanism to focus on the most relevant patches within a bag while simultaneously imposing clustering constraints to ensure that similar patches receive comparable attention weights, enhancing interpretability and performance. We adapted the CLAM algorithm to further improve generalizability by only showing a random subset of patches of a WSI (usually 5%) to the model during an epoch. Moreover, a cosine annealing learning rate scheduler as well as gradient accumulation were added. The Chowder architecture employs a modular and hierarchical approach to MIL, where instance-level features are aggregated through a series of convolutional and pooling layers to capture complex patterns and dependencies. It also incorporates importance sampling to dynamically select the most informative instances from each bag, thereby reducing computational load and enhancing the model’s ability to focus on critical data points.

#### Data Representation

Patches of chromogenic samples were encoded as any other natural image as RGB images (3 color channels) similar to natural images. In contrast, multiplex fluorescent immunostainings required further preprocessing, since in these cases each channel contains the intensity values of one of the respective stainings which does not coincide with color channels. Each of the fluorescence WSIs has 4 channels, containing 3 of the stainingset-specific stainings as well as DAPI, as nuclear (DNA) label. We tested 3 different data representations on the fluorescence WSIs:

- **RGB** This representation treats each of the 3 stainings as RGB color channels and additionally overlays the DAPI channel as a grayscale image on each of the other 3 channels, resulting in an RGB image similar to what can be seen with a standard visualization software when showing all stainings simultaneously. This representation contains the co-occurrence of the stainings, i.e. the model can learn which stainings occur together in the WSI.
- **Channel-wise** The channel-wise representation treats each channel (staining) individually as a grayscale image. Consequently, each patch gets encoded 4 times during feature extraction. The resulting features (1-dimensional vectors) are then appended, leading to a representation where each channel is encoded in detail, but no co-occurrence information is preserved.
- **Channel-wise average** The channel-wise average representation is very similar to the channel-wise representation but aims to include the co-occurrence of stainings by averaging the 4 different feature vectors respective to their occurrence in the image while losing specific details of individual stainings.

#### Training Procedure

We split the data consisting of 100 unique patients into a train (75 patients) and test (25 patients) set and performed the model development solely on the train data. For that, we used a 15-fold cross-validation, where in each fold we randomly used 60 patients from the train set for training and the remaining 15 patients for validation. Due to the different distributions of T2D within the two cohorts, we applied balanced sampling during training only for Tübingen patients while the Dresden cohort was already balanced.

As the target metric, we chose the Area Under the Receiver Operating Characteristic Curve (AUROC), since for the subsequent XAI steps it is more important to have a model that has a high separability and can generally distinguish between the two classes (T2D or not) than having a model that works well with a specific cutoff as measured by the F1-Score or the Accuracy.

### Explainable AI

Attention Methods. Initially, we focused on determining the significance of specific patches using the built-in attention mechanism of the classification head. Nevertheless, raw attention data predominantly indicates general importance rather than class-specific relevance. To address the latter, we applied Attention Layer-wise Relevance Propagation (Attention LRP) (Chefer et al., 2021), which filters the attention values to highlight patches distinctly associated with either diabetic or non-diabetic status within the same WSI. The class-specific relevance R is iteratively computed for each layer n. Here x^(n)^ is the input at layer n with j elements and x^(n-1)^ is the input of the downstream layer with i elements. w_ji_ is the respective weight matrix of the layer. Non-linear functions like GELU (Hendrycks & Gimpel, 2023) produce both positive and negative outputs. To account for this, relevance propagation can be adjusted by forming a subset of indices q resulting in the following relevance propagation:

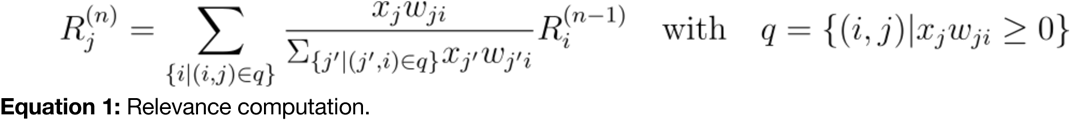

We set R^(0)^=1_t_ for initialization, where *t* is the one-hot-encoded outcome class. Based on the relevance, we compute the average relevance filtered attention *Ā* for each attention head *b*, with the number of head *h*. To this end, we take the Hadamard product between the relevance and the attention’s heads gradient *∇A^(b)^*, considering only the positive relevance:

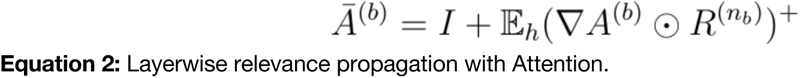

#### Attribution Methods

Compared to attention methods, attribution methods are architecture agnostic and do not require attention mechanisms in the model. The simplest form of attribution is the gradient from an outcome class *y_j_* w.r.t. the input *x.* When only considering the positive gradients, this method is called Saliency (Simonyan et al., 2013). However, raw gradients are prone to gradient shattering (Balduzzi et al., 2017) and vanishing (Bengio et al., 1994). Therefore, we employed Integrated Gradients (IG) (Sundararajan et al., 2017), a more robust technique that offers greater contextual information for each input pixel by incorporating gradients with respect to a baseline value (we chose a black pixel as the baseline). To this end, we compute a path integral between a baseline value *x_0_* and the true value *x_j_* of each of the *j* input pixels:

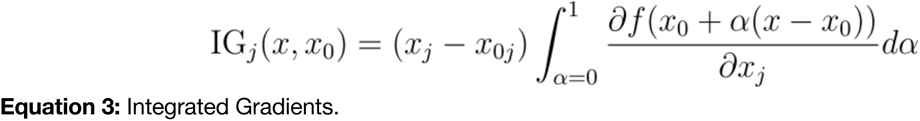

However, the initial selection of a baseline value in IG can be ambiguous, and computing multiple path integrals across various baseline values may be inefficient. To address this, Expected Gradients (EG) (Erion et al., 2020) circumvents the need for baseline selection by utilizing a probabilistic baseline *D* derived from a sample of observations:

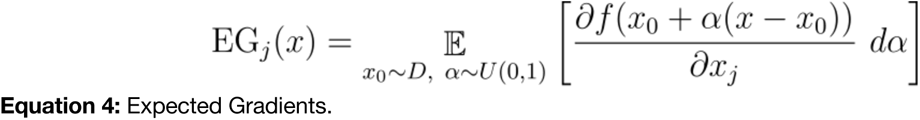

In practice, this expectation is approximated using a mini-batch sampling method for *x_0_* and *α*. More related to relevance computing methods, DeepLIFT (Shrikumar et al., 2017) works by comparing the activation of each neuron to its ’reference activation’ and assigning contribution scores based on the difference. Defining the difference between a target neuron *t* and the baseline *t_0_* as *Δt=t−t_0_* (we chose *t_0_=0*), the summation-to-delta of contribution scores is established. Here, *η* represents the downstream hidden layer neuron, *K,* the number of downstream neurons, and *C,* the single contribution score, which quantifies the impact of downstream neuron *η_i_* on the difference in *t*. Based on the contribution scores, the multiplier *m_ΔηΔt_* is constructed, inspired by the partial derivative in the backpropagation algorithm:

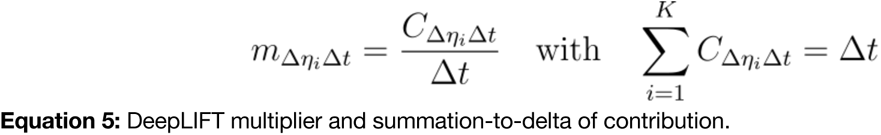

Based on this multiplier, we measure the contribution scores of the input: *m_ΔxΔt_*. To make results more robust, we advance all attribution methods by SmoothGrad (Smilkov et al., ^2^017). SmoothGrad computes the heatmaps *n_SG_* times based on noisy input samples with Gaussian noise (with *σ=1.0* and *n_SG_=10* in our case) and subsequently averages the heatmaps. The heatmaps are smoothed with a Gaussian Kernel with *σ=1.0.* We focus exclusively on positive attribution, emphasizing factors that support a specific outcome. Exemplary heatmaps of all methods are depicted in Suppl. Fig. 9.

### Histologic Biomarker

#### ROI Segmentation

To compute the identified biomarkers a segmentation map of the respective regions is needed. Due to the amount and size of the WSIs we used scribble annotation (Gotkowski et al., 2024) to efficiently label 44 (out of 600 chromogenic WSIs) slides with the labels background, tissue, adipocytes, fibrotic patterns, and islets. Scribble annotation allows for a very fast annotation because regions are not annotated densely (i.e. each pixel gets annotated) but rather only smaller lines (scribbles) are used to approximately label the respective regions. During model training, we used the ignore label in nnU-Net (Isensee et al., 2021) to only update the model weights based on annotated scribbles while ignoring the remaining image. While annotation is based on scribbles, the model still returns a dense prediction which makes this approach well-suited for segmenting the ROIs. Since the nature of scribbles only allows a quantitative evaluation of the respective annotated areas, a qualitative analysis of the resulting segmentation maps was conducted. After adjusting some annotations and retraining the nnU-Net, a satisfactory segmentation quality was reached.

#### Quantifying Attention

The attention to specific ROIs for the chromogenic stainings was computed by summing up the raw attention within specific segmentation masks and standardizing it by the total attention in the WSI and the tissue size. We summed up the attention per staining channel for the fluorescence stainings and standardized it by the total attention overall stainings and the tissue size. For the exploratory analysis, we z-standardized all continuous variables on non-interpretable scales; however, we routinely z-standardized all continuous covariates for the statistical analysis.

#### Biomarker

For the chromogenic staining, we computed the area-related histologic biomarkers based on the segmentation masks and counted the number of distinct islets and adipocyte clusters by applying connected components to the respective segmentation mask. All biomarkers are standardized by the total size of the tissue. For the islet-related biomarkers, we only included islet segmentations larger than 5000 pixels, ignoring segmentation artifacts or errors. For the fluorescence-specific intensity biomarkers, we computed the channel-specific mean intensity as an approximation of the respective area of the trait and standardized it again by the tissue size. In the case of perilipin 1, we first threshold the intensity values, including only values larger than 1000 (intensity measured as 16-bit integers), filtering out unspecific stained traits.

### Statistical Methods

#### Hypothesis Testing

To test differences between two groups, e.g. patients with and without diabetes, we applied a two-sample t-test. To test if at least one group is significantly different, we applied the F-test within a one-way ANOVA. To test if a coefficient in a linear model is significantly different from zero we used the Z-test. For all tests, we considered p-value > as not significant.

Statistical Models. For statistical analysis of the biomarkers, we used a generalized mixed linear model (GMLM), as we have several dependence structures within the data. For the diabetes status as the outcome, we selected the canonical binomial link function. For the HOMA2B level, we selected the canonical Gaussian and Gamma link functions and reported results for the Gaussian link, as its Akaike Information Criterion (AIC) was lower. We accounted for random effects linked to the cohort and staining method. Due to no variation between the outcomes (i.e., diabetes status) within a person, we could not model the person as a random effect. In this case, the intercept would be enough to fit one model per patient perfectly. All models were fitted via maximum likelihood and the L-BFGS-B optimizer (Zhu et al., 1997). We tested for multicollinearity by computing variance inflation factor (VIF) and removed “benign tumor” due to perfect multicollinearity with “malignant tumor” combined with “chronic pancreatitis”.

#### Statistical Model Evaluation

For all models with the same modality and response variable, we used the AIC for model selection. Additionally, we evaluated the logistic models through a confusion matrix and the linear models through parity plots, residual plots, and Q–Q plots of the residuals. All evaluation results of the statistical models are shown in Suppl. Figures 10 & 11.

## Data Availability

The data is publicly available at: https://syncandshare.desy.de/index.php/s/AH3o9jmtNELWtgm. After acceptance the data will be moved to an open repository that adheres to the FAIR data principles (e.g. Zenodo).

## Acknowledgments

1. We thank all the participants of the “Studying Islets from Living Donors” programs in Tübingen and Dresden. The studies were supported by the German Center for Diabetes Research (Deutsches Zentrum für Diabetesforschung, DZD). The DZD is funded by the German Federal Ministry for Education and Research and the states where its partner institutions are located (01GI0925).
2. The authors acknowledge the project specific financial support of the Helmholtz Association (project DIADEM, ZT-1-PF-5 139).
3. This project has received funding from the European Union’s Horizon Europe research and innovation program under grant agreement **No 1010954433 (Intercept-T2D)**. Views and opinions expressed are however those of the author(s) only and do not necessarily reflect those of the European Union. Neither the European Union nor the granting authority can be held responsible for them.
4. Special thanks to Rebekka Wehner, Susanne Doms, and Marc Schmitz (Institute for Immunology of the University Hospital Carl Gustav Carus Dresden) for supporting the development of the multiplex immunofluorescence staining.
5. This work was funded by Helmholtz Imaging (HI), a platform of the Helmholtz Incubator on Information and Data Science.
6. Birkenfeld A. was supported by the German Federal Ministry for Education and Research (01GI0925) via the German Center for Diabetes Research (DZD eV); Ministry of Science, Research and the Arts Baden-Württemberg; and Helmholtz Munich.
7. The authors thank Darya Trofimova, Lars Krämer and Carsten Lüth of the DKFZ for insightful discussions and feedback.
8. This work was supported by the Light Microscopy Facility, a Core Facility of the CMCB Technology Platform at TU Dresden.

