## supplementary_material for "Explainable AI-based analysis of human pancreas sections identifies traits of type 2 diabetes"

### Appendix

#### Supplementary Figures

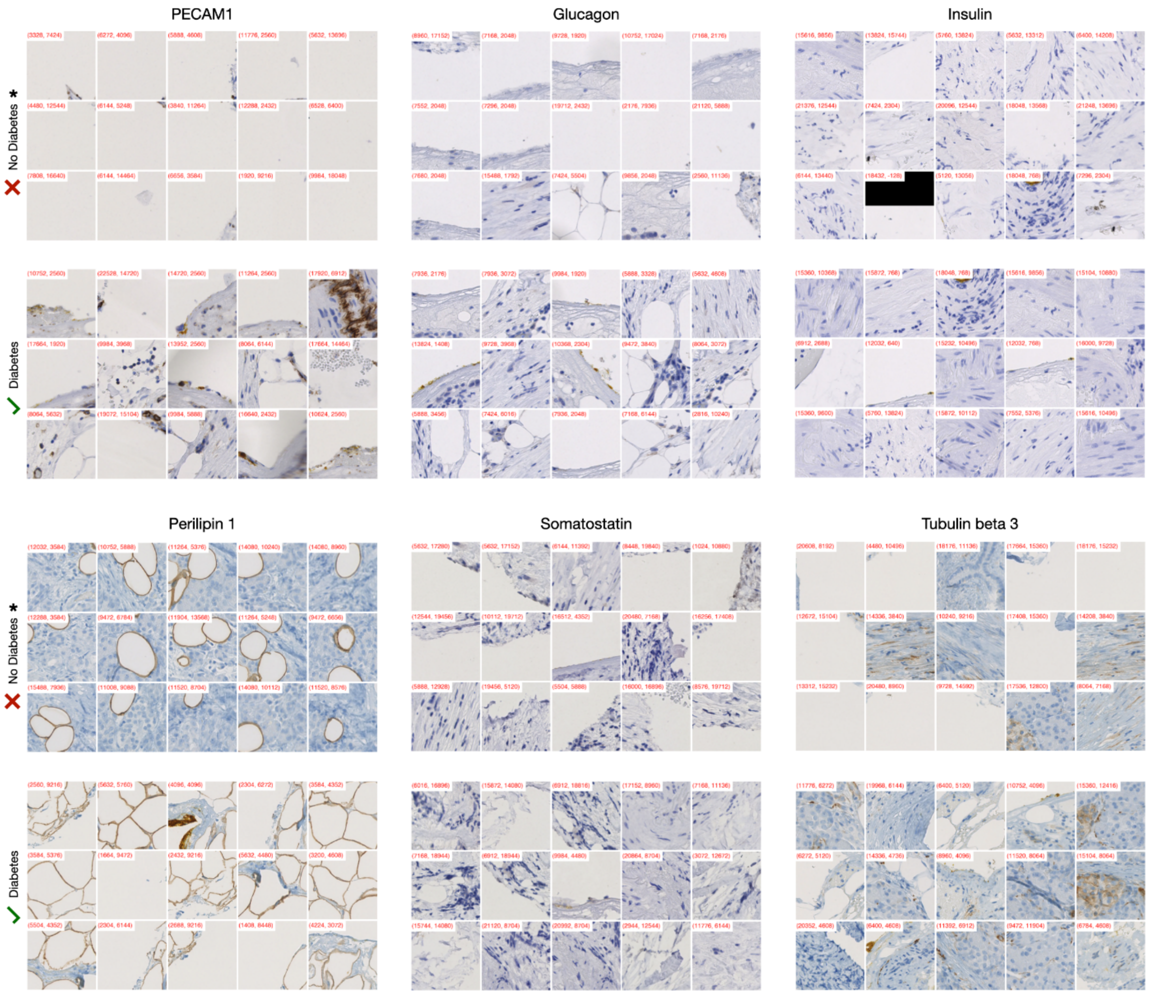


**Suppl. Figure 1:** Representative top 15 attended patches from each chromogenic staining associated with the outcomes ‘diabetes’ and ‘no diabetes’.

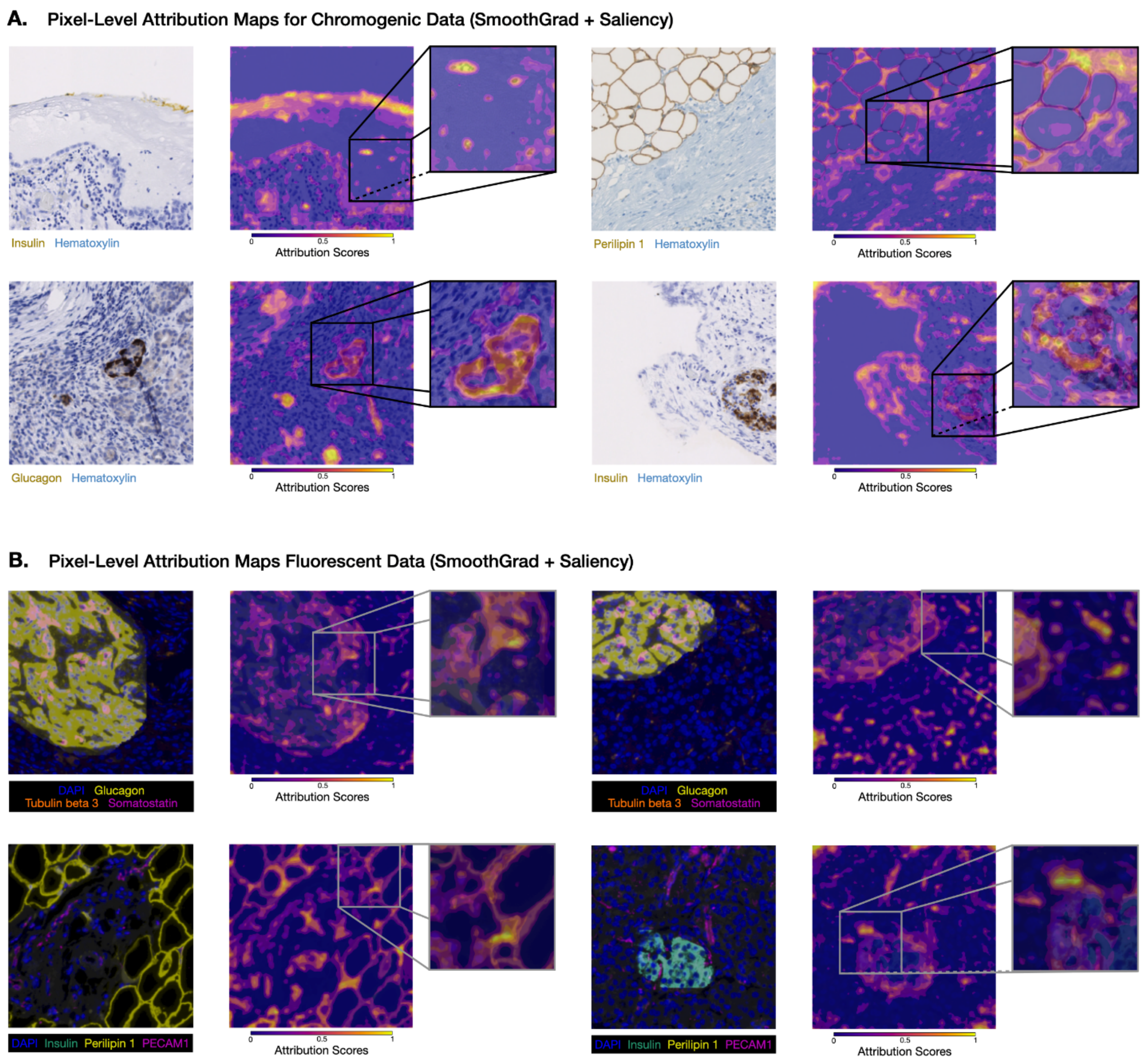


**Suppl. Figure 2:** Pixel-level heatmaps of local regions and one single patch based on Saliency with SmoothGrad for chromogenic (**A**) and fluorescence (**B**) data.

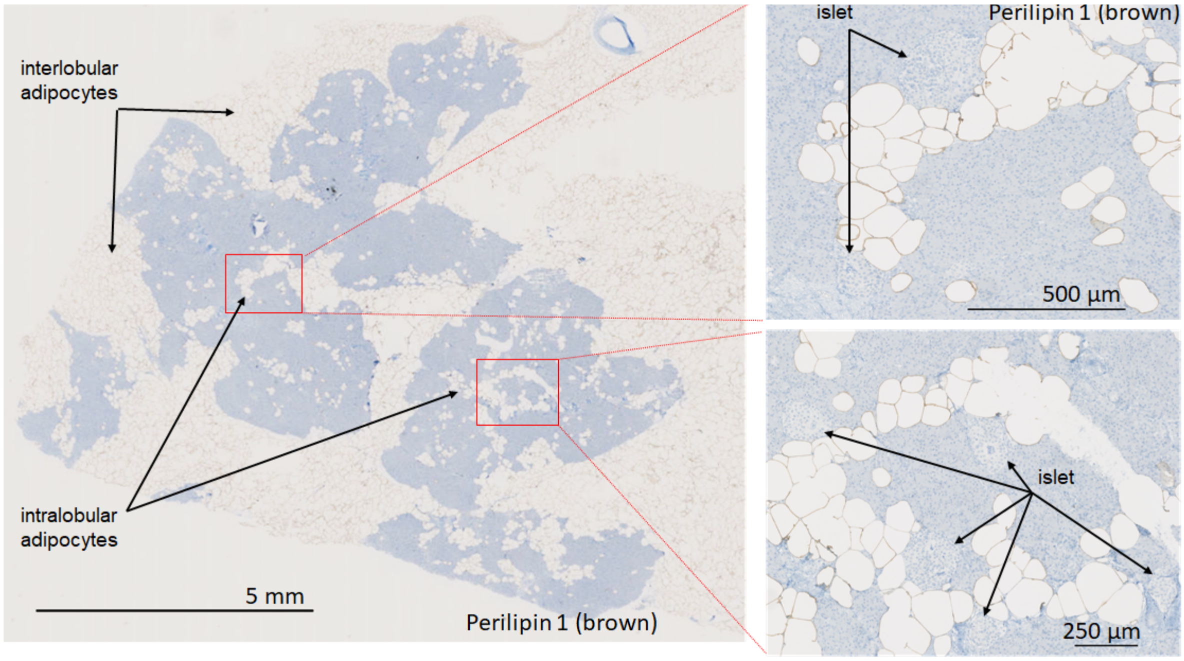

**Suppl. Figure 3:** Representative brightfield image (zoom-in) of a pancreatic section from a non-diabetic patient stained for the adipocyte marker perilipin 1 (brown), showing inter- and intra-lobular infiltration of pancreatic parenchyma with adipocytes. Note the close proximity between adipocytes and some of the islets.

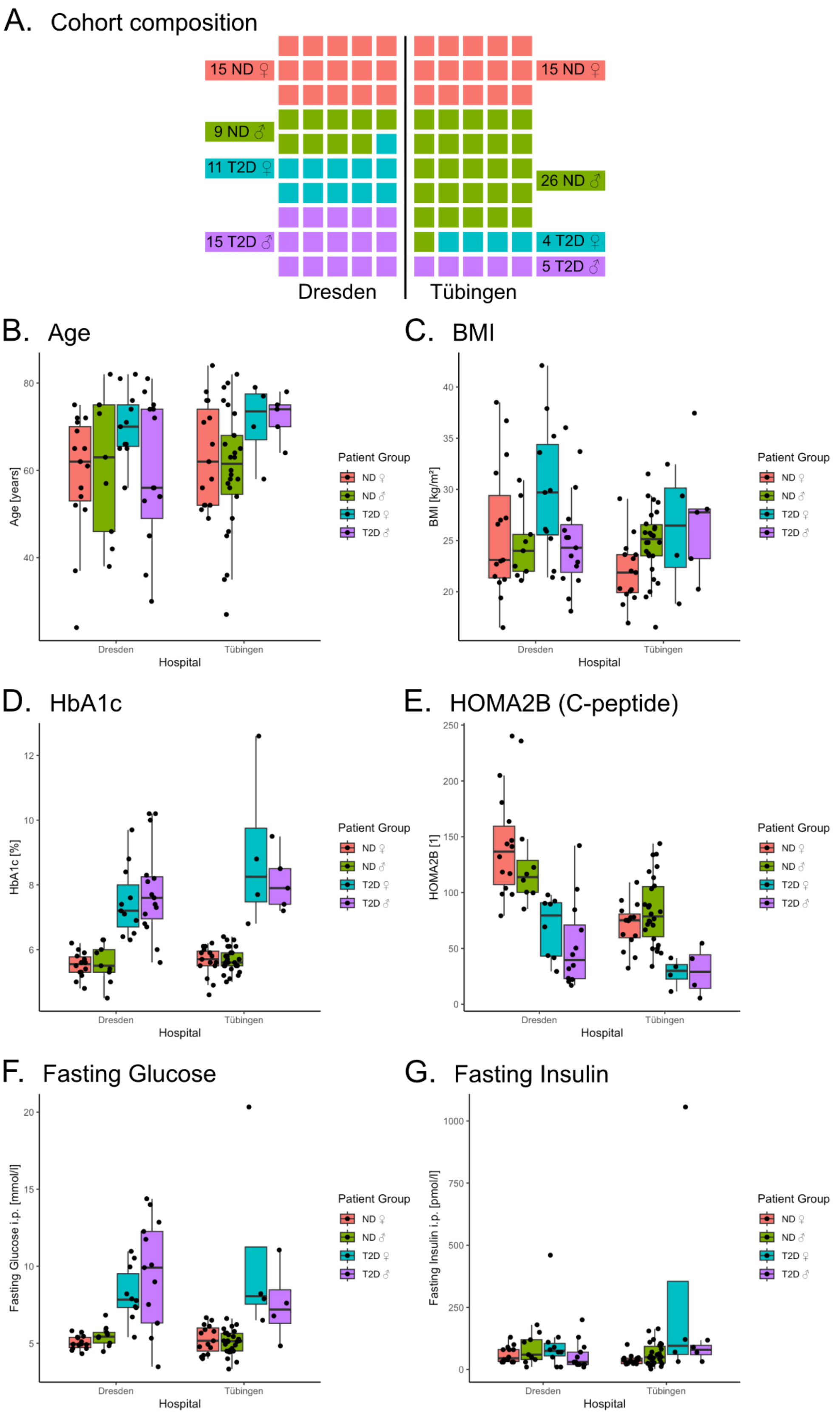


**Suppl. Figure 4:** Composition and selected clinical data of the analyzed cohort**.** 100 pancreatectomized patients were recruited for this study, including female (♀) and male (♂) individuals with non-diabetic (ND) or type 2 diabetic (T2D) background. 50 patients each were treated due to different indications (e.g. pancreas carcinoma or pancreatitis) at the University Hospitals in Dresden and Tübingen (**A**). T2D patients were diagnosed as such according to their clinical records minimum of one year before the onset of the pancreatic indication. All ND patients do not fulfill the diagnostic criteria for T2D defined by the American Diabetes Association (ADA). Clinical data regarding age (**B**), body weight via body mass index (BMI, **C**), long-term hyperglycemia via glycated hemoglobin (HbA1c, **D**), β-cell function, and insulin resistance via homeostasis model assessment 2B including c-peptide (HOMA2B, **E**), fasting glucose (**F**), and fasting insulin (**G**) are showed.

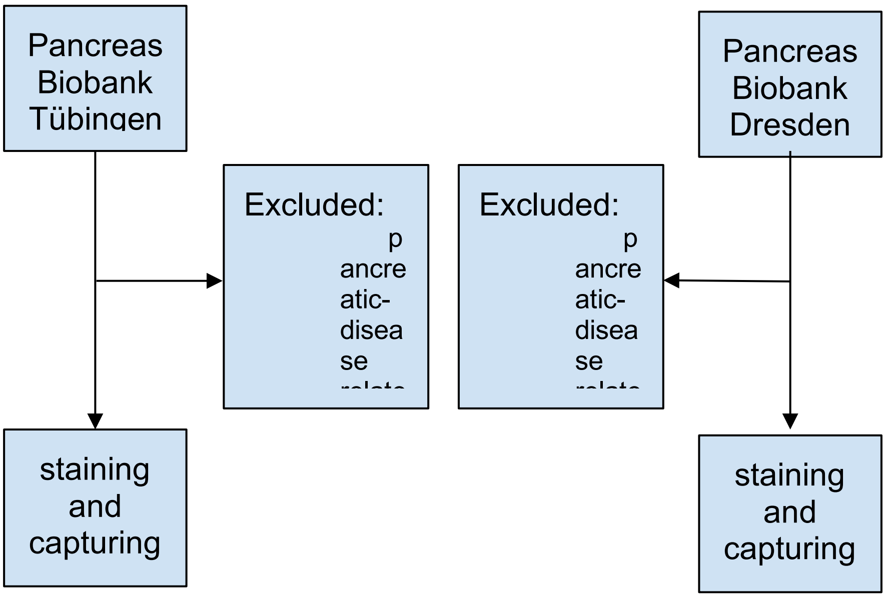


**Suppl. Figure 5:** Sample Selection Process.

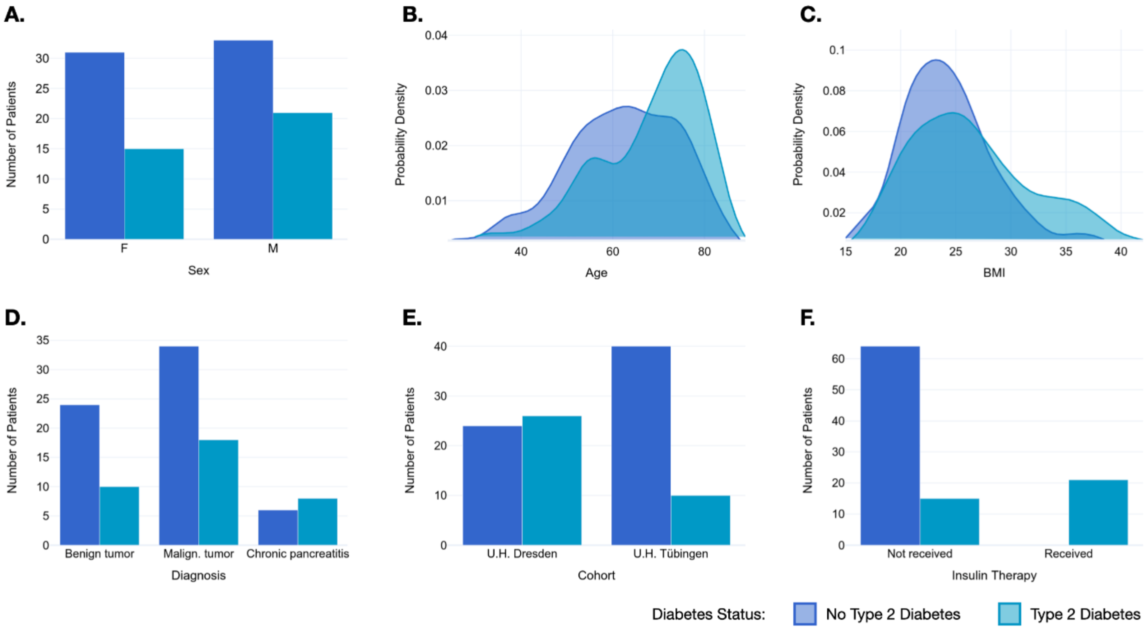

**Suppl. Figure 6:** Distribution of patient clinical data. Distribution of all control variables between both glycaemic statuses, i.e. no diabetes vs T2D, excluding the immunohistological stainings: sex (**A**), age (**B**), BMI (**C**), condition underlying pancreatic surgery (**D**), cohort (**E**), and insulin therapy (**F**).

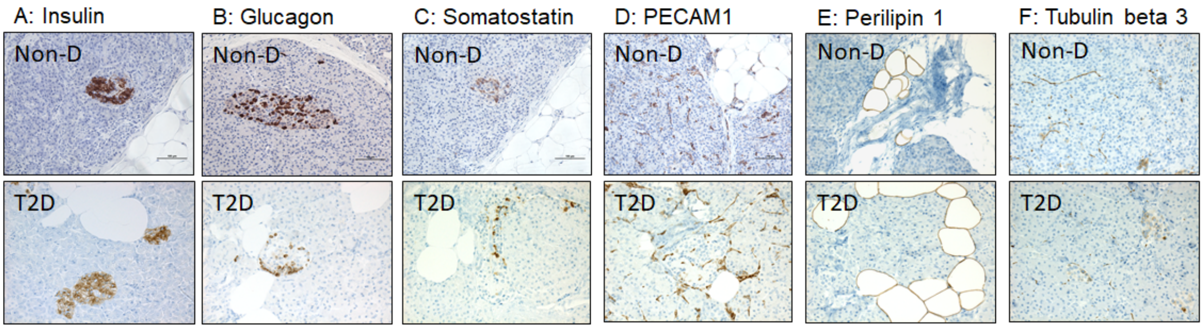


**Suppl. Figure 7:** Representative brightfield microscopy images (20x magnification) of pancreatic resections from a normal glucose-tolerant patient (A-F: upper panels) and a patient with T2D (A-F: lower panels). Sections of formalin-fixed, paraffin-embedded human pancreatic tissue were stained (in brown) for (A) insulin, (B) glucagon, (C) somatostatin, (D) PECAM1, (E) perilipin 1, (F) tubulin beta 3, and counterstained with hematoxylin (blue) as described in Methods.

## A. No T2D

Stainingset 1:

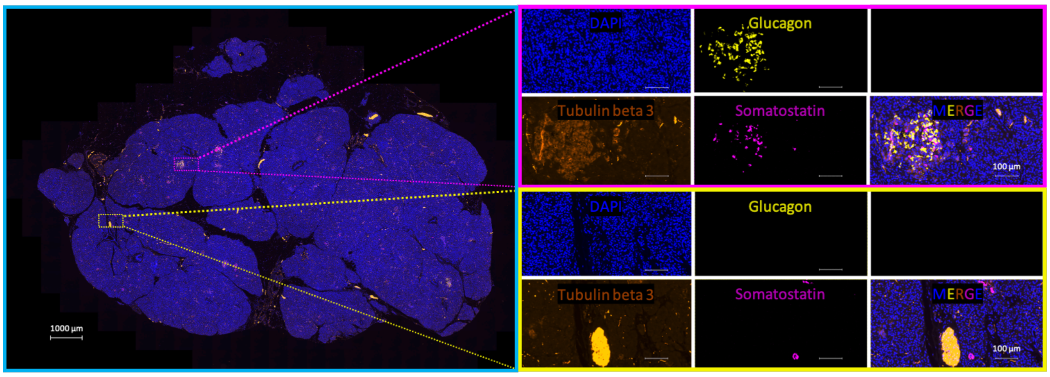


Stainingset 2:

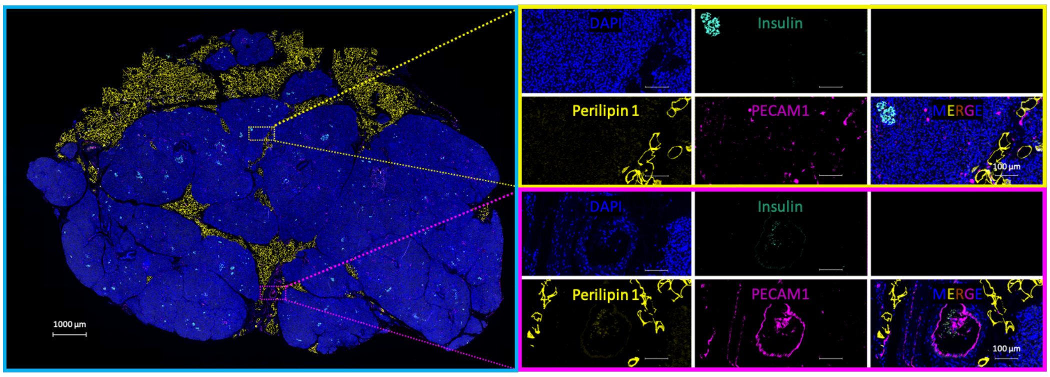


**B. T2D**

---

Stainingset 1:

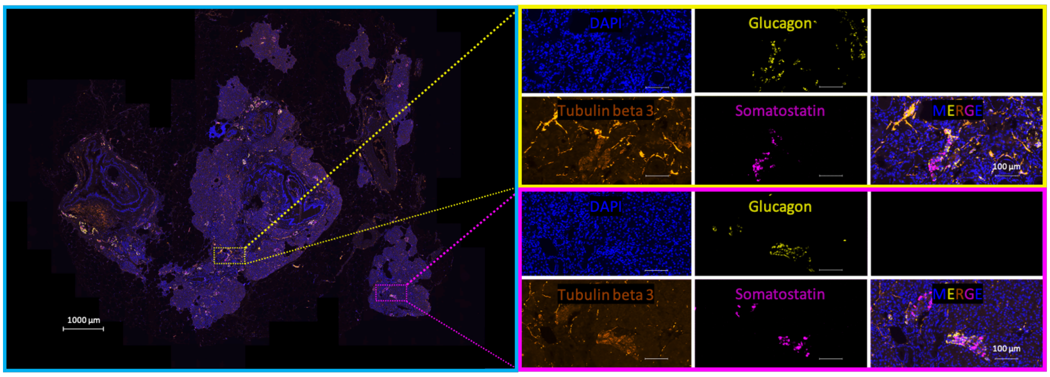


Stainingset 2:

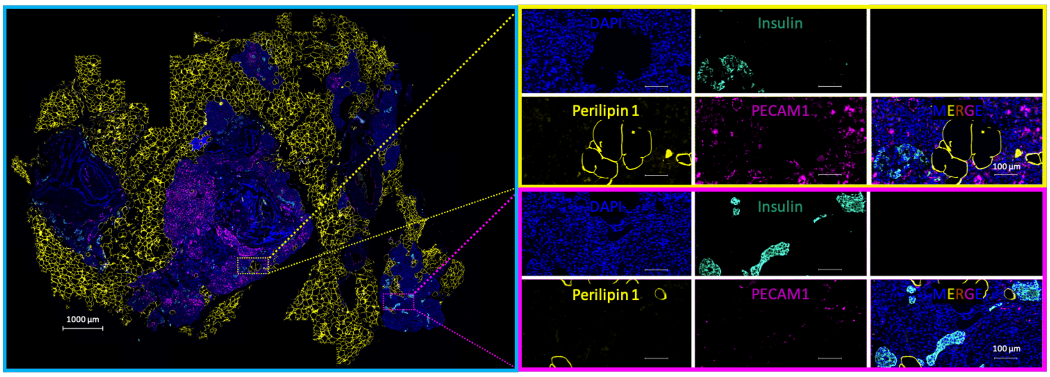


**Suppl. Figure 8:** Representative fluorescence microscopy images of pancreatic resections from the same normoglycemic patient (**A**) and the same patient with type 2 diabetes (**B**) as showcased for chromogenic stainings in Suppl. Figure 7. Nuclear DNA is visualized using DAPI. Stainingset 1: glucagon and somatostatin are markers for pancreatic α-and δ-cells, respectively whereas tubulin beta 3 marks neuronal axons (intense fluorescence) as well as islet cells (moderate fluorescence). Stainingset 2: insulin, perilipin 1, and PECAM1 are markers for pancreatic β-cells, adipocytes, and vascular endothelial cells, respectively. Whole-slide images were captured as tile scans with 20x magnification. Overview images's scale bar: 1000 µm; detail image's scale bar: 100 µm.

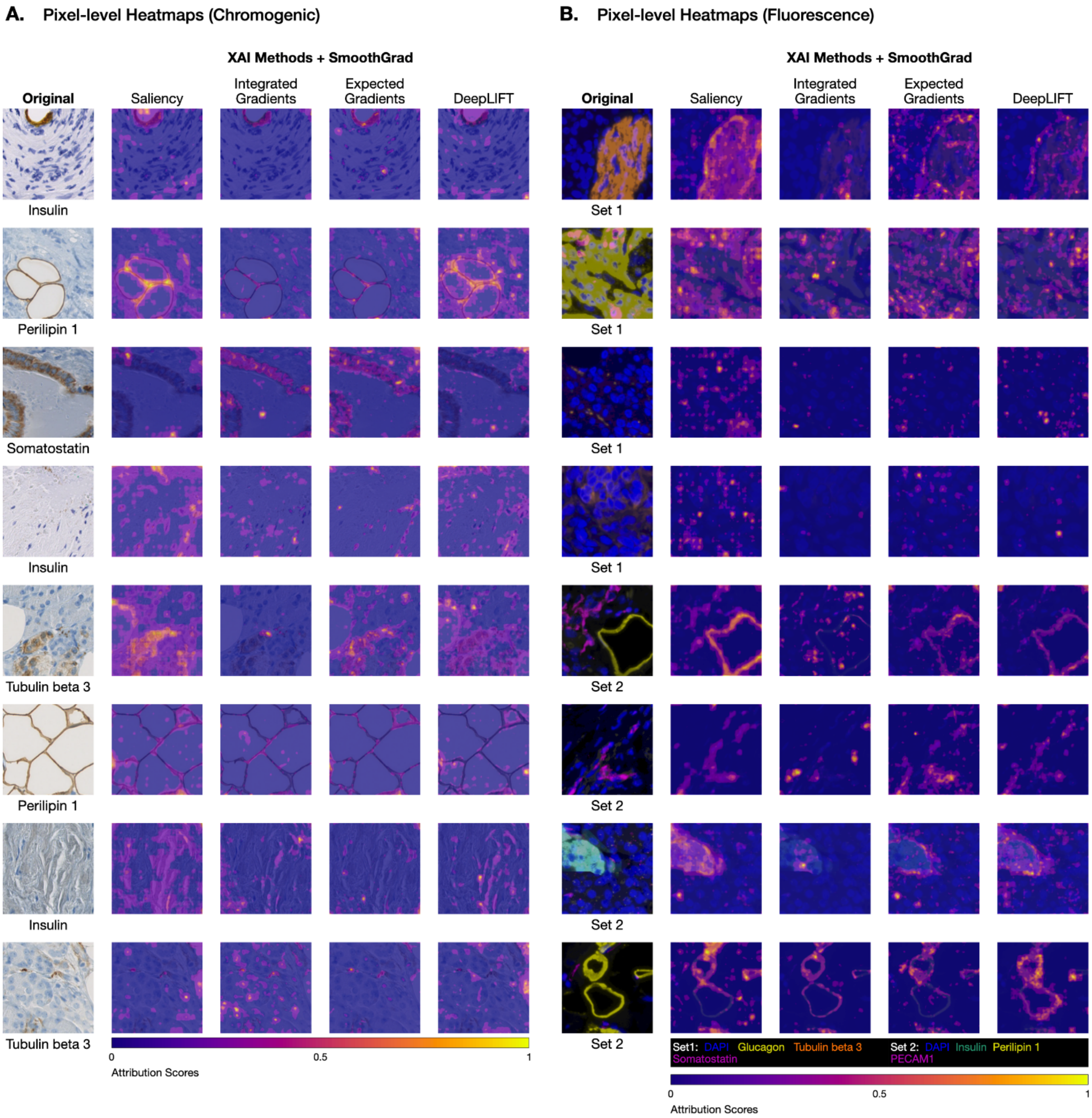


**Suppl. Figure 9:** Exemplary pixel-level heatmaps of all four applied attribution methods for chromogenic (**A**) and fluorescently (**B**) stained patches. They differ in granularity and in rare occasions also in attributed regions.

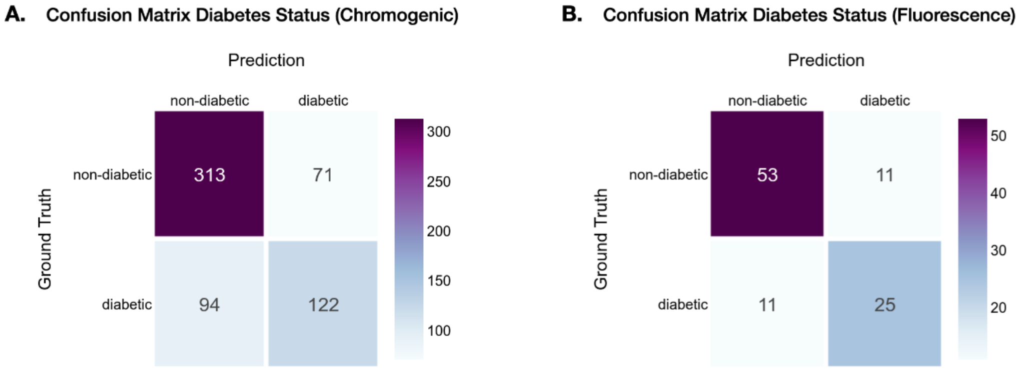


**Suppl. Figure 10:** Logistic MLM confusion matrices.

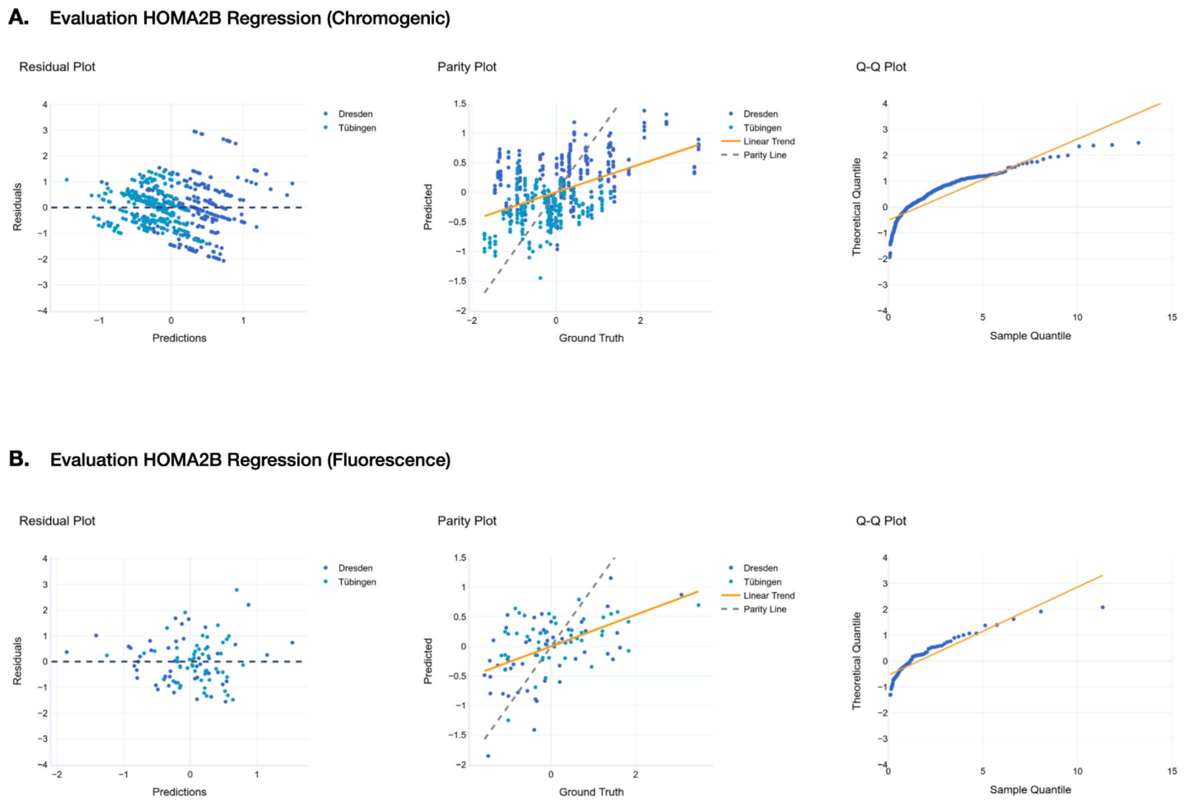


**Suppl. Figure 11:** Residual, Parity, and Q-Q Plot for HOMA2-B predicting MLMs on chromogenic (**A**) and fluorescence (**B**) data.

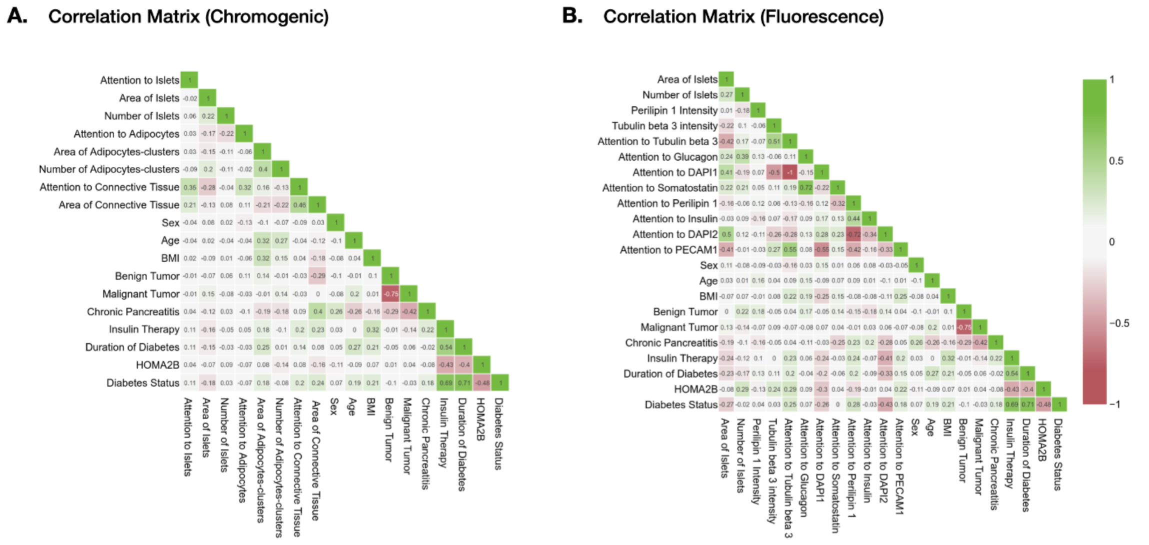


**Suppl. Figure 12:** Correlation between biomarker and clinical patient data for chromogenic (**A**) and fluorescence (**B**) data sets.

### Supplementary Tables

##### A: Ensemble Performance on Chromogenic WSIs

| Representation | Stainingset | Encoder / MIL Algorithm | AUROC (higher is better) |
|---|---|---|---|
| Channel-wise Average | 1 | Imagenet / CLAM | **0.956** |
| | 2 | Imagenet / CLAM | 0.684 |
| Channel-wise | 1 | Imagenet / CLAM | **0.912** |
| | 2 | Imagenet / CLAM | 0.816 |
| RGB | 1 | Imagenet / CLAM | **0.842** |
| | 2 | Imagenet / CLAM | 0.640 |

**B: Ensemble Performance on Fluorescence WSIs**

| Staining | MIL Algorithm | Encoder | AUROC (higher is better) |
|---|---|---|---|
| Mean over Stainings | CLAM | Imagenet | **0.833** |
| | Chowder | Imagenet | 0.830 |
| | CLAM | Phikon | 0.794 |
| | Chowder | Phikon | 0.781 |
| Tubulin beta 3 | CLAM | Imagenet | **0.895** |
| Insulin | CLAM | Imagenet | 0.842 |
| Glucagon | CLAM | Imagenet | 0.842 |
| Perilipin 1 | CLAM | Imagenet | 0.842 |
| Somatostatin | CLAM | Imagenet | 0.842 |
| PECAM1 | CLAM | Imagenet | 0.737 |
| Tubulin beta 3 | CLAM | Phikon | 0.851 |
| Insulin | CLAM | Phikon | 0.684 |
| Glucagon | CLAM | Phikon | 0.711 |
| Perilipin 1 | CLAM | Phikon | 0.816 |
| Somatostatin | CLAM | Phikon | **0.921** |
| PECAM1 | CLAM | Phikon | 0.781 |
| Tubulin beta 3 | Chowder | Imagenet | **0.877** |
| Insulin | Chowder | Imagenet | 0.851 |
| Glucagon | Chowder | Imagenet | 0.640 |
| Perilipin 1 | Chowder | Imagenet | **0.877** |
| Somatostatin | Chowder | Imagenet | **0.877** |
| PECAM1 | Chowder | Imagenet | 0.851 |
| Tubulin beta 3 | Chowder | Phikon | 0.833 |
| Insulin | Chowder | Phikon | 0.693 |
| Glucagon | Chowder | Phikon | 0.728 |
| Perilipin 1 | Chowder | Phikon | **0.860** |
| Somatostatin | Chowder | Phikon | 0.842 |
| PECAM1 | Chowder | Phikon | 0.728 |

**Suppl. Table 1:** Ensemble AUROC for prediction performance of each model for the chromogenic (A) and fluorescence (B) WSIs on the held-out test set consisting of 25 patients.

| | Tübingen (% or mean±SD) | Dresden (% or mean±SD) |
|---|---|---|

| | | |
|---|---|---|
| Sex (females) | 38 % | 52 % |
| Age (years) | 63.2 ± 12.97 | 61.8 ± 14.35 |
| BMI (kg/m2) | 26.33 ± 5.8 | 24.30 ± 4.1 |
| Diabetes (%) | 18 | 52 |
| - diabetes duration (years) | 2.2 ± 6.5<br>(0-35 years) | 5.7 ± 8.8<br>(0-30 years) |
| - metformin treatment | 6 (12 %) | 11 (22%) |
| - sulfonylurea treatment | 1 (2%) | 2 (4%) |
| - GLP-1 analogon treatment | 0 (0 %) | 3 (6%) |
| - SGLT2-inhibitor treatment | 0 (0 %) | 1 (2 %) |
| - insulin treatment | 6 (12 %) | 15 (30%) |
| Fasting glucose (mmol/l) | 5.8 ± 2.49 | 7.07 ± 2.78 |
| Fasting insulin (pmol/l) | 76.80 ± 147.8 | 74.35 ± 74.08 |
| HOMA-2B (C-peptide) | 70.76 ± 32.15 | 99.58 ± 55.56 |
| HbA1c (%) | 6.17 ± 1.35 | 6.67 ± 1.44 |
| Diagnosis | | |
| - Malignant disease | 58 % | 46 % |
| - Chronic Pancreatitis | 6 % (3 out of 50) | 22 % (11 out of 50) |
| - Other | 36 % | 32 % |

**Suppl. Table 2:** Clinical cohort characteristics.

**A. GMLM Regression for Diabetes Status (Fluorescence)**

| Fixed Effects | Estimate | SE | P-Value *(p)* | Sign. |
|---|---|---|---|---|
| Intercept | -1.889 | 0.877 | 0.031 | * |
| Area of Islets | -0.446 | 0.426 | 0.295 | |
| Number of Islets | 0.085 | 0.288 | 0.768 | |
| Perilipin 1 Intensity | 0.408 | 0.265 | 0.124 | |
| Tubulin beta 3 Intensity in Islets | -0.328 | 0.310 | 0.168 | |
| Sex | 0.709 | 0.554 | 0.201 | |
| Age | 0.832 | 0.330 | 0.012 | * |
| BMI | 0.593 | 0.263 | 0.024 | * |
| Malignant Tumor | 0.672 | 0.616 | 0.276 | |
| Chronic Pancreatitis | 1.915 | 0.967 | 0.048 | * |

| Random Effects | Type | Variance | STD |
|---|---|---|---|
| Cohort | Intercept | 0.754 | 0.868 |
| Observations | 100 | | |
| Groups | {Cohort: 2} | | |
| Log-likelihood | -51.628 | | |
| AIC | 125.256 | | |
| *Note* | ** p<0.05, ** p<0.01, *** p<0.001* | | |

**B. MLM Regression for HOMA2B (Fluorescence)**

| Fixed Effects | Estimate | SE | P-Value *(p)* | Sign. |
|---|---|---|---|---|
| Intercept | 0.306 | 0.173 | 0.080 | |
| Area of Islets | -0.207 | 0.094 | 0.030 | * |
| Number of Islets | 0.280 | 0.103 | 0.008 | ** |
| Perilipin 1 Intensity | 0.032 | 0.085 | 0.706 | |
| Tubulin beta 3 Intensity in Islets | 0.183 | 0.093 | 0.053 | |
| Sex | -0.149 | 0.170 | 0.383 | |
| Age | -0.154 | 0.088 | 0.085 | |
| BMI | 0.243 | 0.090 | 0.008 | ** |
| Malignant Tumor | 0.076 | 0.187 | 0.685 | |
| Chronic Pancreatitis | 0.314 | 0.307 | 0.309 | |
| Insulin Therapy | -1.435 | 0.244 | 0.000 | *** |

| Random Effects | Type | Variance | STD |
|---|---|---|---|
| Cohort | Intercept | 0.000 | 0.000 |
| Observations | 91 | | |
| Groups | {Cohort: 2} | | |
| Log-likelihood | -105.898 | | |
| AIC | 237.797 | | |
| *Note* | ** p<0.05, ** p<0.01, *** p<0.001* | | |

**Suppl. Table 3:** Regression results of the generalized mixed linear model analysis for (A) diabetes status and (B) HOMA2B levels based on fluorescence stainings.
